# Medication information extraction using local large language models

**DOI:** 10.1101/2025.03.28.25324847

**Authors:** Phillip Richter-Pechanski, Marvin Seiferling, Christina Kiriakou, Dominic M. Schwab, Nicolas A. Geis, Christoph Dieterich, Anette Frank

**Author notes:** Corresponding author, CD and AF contributed equally.

## Abstract

Medication information is crucial for clinical routine and research. However, a vast amount is stored in unstructured text, such as doctoral letters, requiring manual extraction -- a resource-intensive, error-prone task. Automating this process comes with significant constraints in a clinical setup, including the demand for clinical expertise, limited time-resources, restricted IT infrastructure, and the demand for transparent predictions. Recent advances in generative large language models (LLMs) and parameter-efficient fine-tuning methods show potential to address these challenges.

We evaluated local LLMs for end-to-end extraction of medication information, combining named entity recognition and relation extraction. We used format-restricting instructions and developed a feedback pipeline to facilitate automated evaluation. We applied token-level Shapley values to visualize and quantify token contributions, to improve transparency of model predictions.

Two open-source LLMs -- one general (Llama) and one domain-specific (OpenBioLLM) – were evaluated on the English n2c2 2018 corpus and the German CARDIO:DE corpus. OpenBioLLM frequently struggled with structured outputs and hallucinations. Fine-tuned Llama models achieved new state-of-the-art results, improving F1-score by 10% for adverse drug events and 6% for medication reasons on English data. On the German dataset, Llama established a new benchmark, outperforming traditional machine learning methods by 16% micro average F1-score.

Our findings show that fine-tuned local open-source generative LLMs outperform SOTA methods for medication information extraction, delivering high performance with limited time and IT resources in a clinical setup, and demonstrate their effectiveness on both English and German data. Applying Shapley values improved prediction transparency, supporting informed clinical decision-making.

**Highlights:**

- *<u>Robust end-to-end medication information extraction with automated evaluation</u>:* We present an end-to-end joint named entity recognition and relation extraction pipeline using generative LLMs, enhanced by an automatic feedback mechanism utilizing a feedback LLM to simplify the oftentimes complex assessment of clinically critical predictions.
- *<u>State-of-the-art performance across languages:</u>* Fine-tuned general LLMs surpassed existing benchmarks by up to 10% in complex medication classes on English data and established a new benchmark for German clinical datasets.
- *<u>Resource-efficient fine-tuning in clinical setup:</u>* We demonstrated that parameter-efficient fine-tuning of local open-source LLMs yields consistent structured outputs and superior extraction performance, addressing clinical constraints like limited expertise, restricted IT infrastructure, and stringent transparency requirements.
- *<u>Enhanced transparency with Shapley values:</u>* We utilized token-level Shapley values tailored for generative LLMs to systematically quantify and visualize individual token contributions, enabling clinicians to better understand and trust model predictions in medication information extraction tasks.

## Introduction

Medication information is crucial in clinical routine, e.g., to ensure safe medication reconciliation during patient admission; to maintain medication continuity for care transitions or to guide life-saving treatment decisions in clinical emergencies. Importantly, medication information is a valuable data source for clinical research.

However, much of medication information is stored in unstructured text, such as doctor’s letters. Figure 1 shows a snippet of a diagnosis section in a doctor’s letter containing medication names and other medication-related information, e.g., duration of a medication and reasons for prescription. Typically, this information must be manually extracted by clinical domain experts, a process which is not scalable, time-consuming and error-prone. Automatic medication information extraction (MIE) methods could facilitate the extraction from large volumes of data, improve data quality, and save valuable time of clinical staff. Recent advances in NLP and machine learning (ML) have demonstrated the potential of generative language models (LLM) and parameter-efficient fine-tuning (PEFT) methods to automate various information extraction tasks.

**Figure 1.**
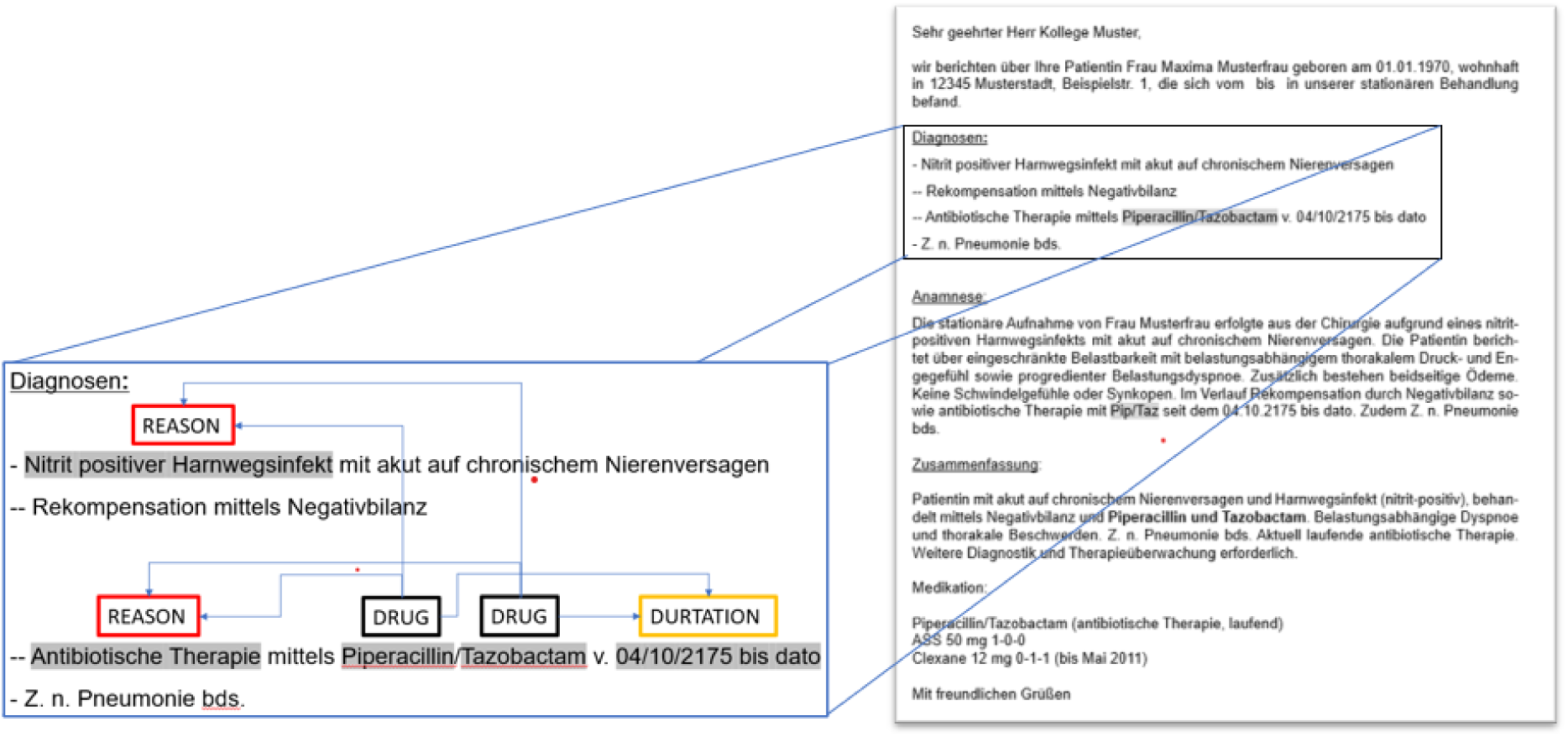
Doctor’s letter: Snippet of a doctor’s letter containing medication information in the diagnosis section.

However, developing MIE methods in a clinical setup comes with four main challenges (Ch).[1], [2]

Ch 1. **Demand of clinical experts** Clinical texts are written in a domain-specific jargon with specific technical terms.[3] These texts need to be manually annotated by clinical experts to prepare them for training and evaluation of ML methods.
Ch 2. **Limited and costly time resources** Due to data protection regulations all experiments need to be conducted by clinical staff in a secure infrastructure. This significantly restricts the availability of personnel and prohibits the involvement of external experts.[4]
Ch 3. **Limited clinical IT resources** Experiments need to be performed inside the clinical IT infrastructure. This excludes the use of scalable external cloud services to access additional CPU and GPU resources.[5]
Ch 4. **Transparent and comprehensive predictions** Clinical routine is a safety-critical domain. Model predictions need to be as transparent and comprehensible as possible.[6]

To address these challenges, we employ state-of-the-art (SOTA) NLP methods, with the following main research contributions:

1. To incorporate clinical domain expertise we utilize the findings from Singhal et al. 2023 claiming that foundation LLMs can encode clinical knowledge.[7] Furthermore, we efficiently evaluate domain-adapted medical LLMs and use clinical knowledge stored in small-sized high-quality annotated gold standard data sets in English and German to fine-tune and evaluate our models yielding new SOTA on difficult evaluation categories.[8] With this, we are the first to evaluate LLMs on a MIE task on clinical routine datasets in two languages (with limited data, and evaluation on gold standard data).
2. To save time resources (1) for pipeline development and maintenance we define our MIE task as a one-step end-to-end (e2e) joint named entity recognition (NER) and relation extraction (RE) task. (2) To support automatic evaluation we use well-defined structured output formats with established evaluation metrics. To address the often-complex process of evaluation of LLMs we utilize feedback LLMs leveraging findings of Chiang et al. 2023. [7], [9], [10] To the best of our knowledge, this is the first study to establish best-practice approaches for structured output generation in a clinical domain for two languages using a feedback LLM for automatic evaluation.
3. Fine-tuning LLMs can be computationally costly, thus we use SOTA PEFT methods based on low-rank adaptation (LoRA), parameter quantization and smaller expert MIE LLMs in a clinical environment.[8], [11]–[13] Thus, we showcase that smaller fine-tuned expert LLMs reliably generate structured output and yield new SOTA results in a MIE task.
4. As previous studies showed, saliency maps can support users in the clinical domain to explain model predictions.[14] To increase transparency and to support our evaluation results we use well-defined Shapley values that are optimized for generative LLMs using the model interpretability library Captum.[15], [16] Finally, our MIE approach extracts structured information from unstructured texts, making the data inherently more interpretable and enabling effective search. Herein we propose a novel experimental setup leveraging saliency maps to support structured MIE evaluation.

### State-of-the-art

#### Generative LLMs

In recent years generative LLMs showed impressive capabilities in various NLP tasks.[17], [18] Google evaluated their large PaLM model for various medical question-answering (QA) tasks, showing that foundation LLMs encode clinical knowledge, thereby underscoring the transformative potential of LLMs in healthcare.[7], [19] Because most SOTA models are closed-source or contain hundreds of billions of parameters, their usage in the resource-restricted and privacy-sensitive clinical routine remains limited. However, the publication of open-source models like Llama and Mistral pushed the development of local LLMs applications and research.[20], [21]

#### Fine-tuning and domain-expert LLMs

Quantization and parameter-efficient fine-tuning methods have enabled further pre-training and fine-tuning of LLMs on constrained IT infrastructure across a range of domains. This has resulted in the release of several medical-domain-adapted LLMs, including PMC-Llama, Meditron or recently OpenBioLLM.[22]–[24] Furthermore, several studies showed that smaller fine-tuned expert LLMs frequently outperform larger general LLMs on various medical tasks.[13], [25], [26]

#### LLMs in the medical domain

While we have seen various studies evaluating LLMs on medical QA data sets (cf. MedQA[27], MedMCQA[28], PubMedQA[29]), works that apply LLMs for MIE tasks remain limited. Steering LLMs to consistently generate structured output is still under intensive research.[30], [31] Recently, encoder-decoder models and generative models have been leveraged to solve NER and RE tasks using well-defined JSON strings as output.[32]–[34] This makes it possible to evaluate these models on existing high-quality benchmark datasets for various IE tasks using well-established metrics, such as precision, recall and F1-score, and to compare their performance to traditional encoder-based classification models. We are the first to use JSON representations in a cardiovascular domain in two languages to represent our one-step e2e MIE task.

#### Medication information extraction

In the context of shared tasks three data sets had been published for the task of medication information extraction (i2b2 2009[35], MADE 1.0 2018[36], n2c2 track 2 2018[37]). All data sets contain named entity annotations and relation annotations. As our English data set, we selected the most recent and comprehensive n2c2 2018 data set. It contains 505 clinical notes extracted from the MIMIC III database [38].

Wei et al, achieved SOTA results for the n2c2 task using a combination of recurrent (RNN) and convolutional neural networks (CNN).[39] El-Allaly et al. used BERT (SciBERT) transformers and established a new SOTA until today.[40] In this study we use both results as our baselines for the English dataset. Most recently some studies used generative LLMs to solve the n2c2 MIE task. Fornasiere et al. compared zero-shot and few-shot performance using Mistral 7b and Hsu et al. developed a weakly-supervised pipeline using Llama-7b and -13b to label clinical data to fine-tune a downstream BERT model. However, neither study reached new SOTA results, and both presented only micro-averaged F1-scores per model, excluding class-wise evaluations.[41], [42] Hu et al. used generative LLMs to investigate the suitability of LLMs for MIE tasks. However, they evaluated their models solely for medication extraction using NER and RE separately without any joint evaluation. Furthermore, they only used a small subset of the i2b2 2010 clinical corpus [43]. Modi et al, in a recent comprehensive survey, present all publications using the n2c2 dataset as a benchmark.[44]

#### Medication information extraction in German

For the German language only a few studies are available. Sharma et al. and Roller et al used a BERT model for medication information extraction. [45], [46] However, their data sets are not publicly available. The only distributable German data set with annotated medication information is currently CARDIO:DE.[47] We selected this as our second benchmark data set. To the best of our knowledge, we are the first to use state-of-the-art generative LLMs on English and German clinical routine data to extract e2e medical information.

#### Interpretability

While LLMs are able to solve various tasks, their black-box character remains challenging, especially in the safety-critical clinical domain.[48] To address such challenges, several feature attribution methods have been developed, especially for classification models.[15], [49], [50] However, assessing their quality is still under active research.[51], [52] To determine the contribution of each input feature to a model prediction, Shapley values provide a theoretically well-founded approach.[53] SHAP is a computationally optimized implementation of Shapley values making them feasible in the clinical domain.[15] To our knowledge, we are the first to study the use of Shapley values for generative LLMs in a clinical domain using a most recent implementation based on Captum.[16]

## Methods

We define our MIE experiments as a one-step e2e joint NER and RE task (graphical abstract: Figure 2). We created an evaluation framework that includes four main steps: (1) data preprocessing (cf. Data preprocessing), (2) fine-tuning an LLM on training data/applying it zero-shot, (3) generating the JSON output (cf. Local large language models), and (4) evaluating the overall performance of the model (using micro average F1-score) and per relation class (using F1-score) (cf. Evaluation and feedback LLM). For inference, we apply (a) data preprocessing, (b) generating the JSON output, and (c) to support interpretability we use saliency maps to visualize input token contributions to the JSON output (cf. Shapley values).

**Figure 2.**
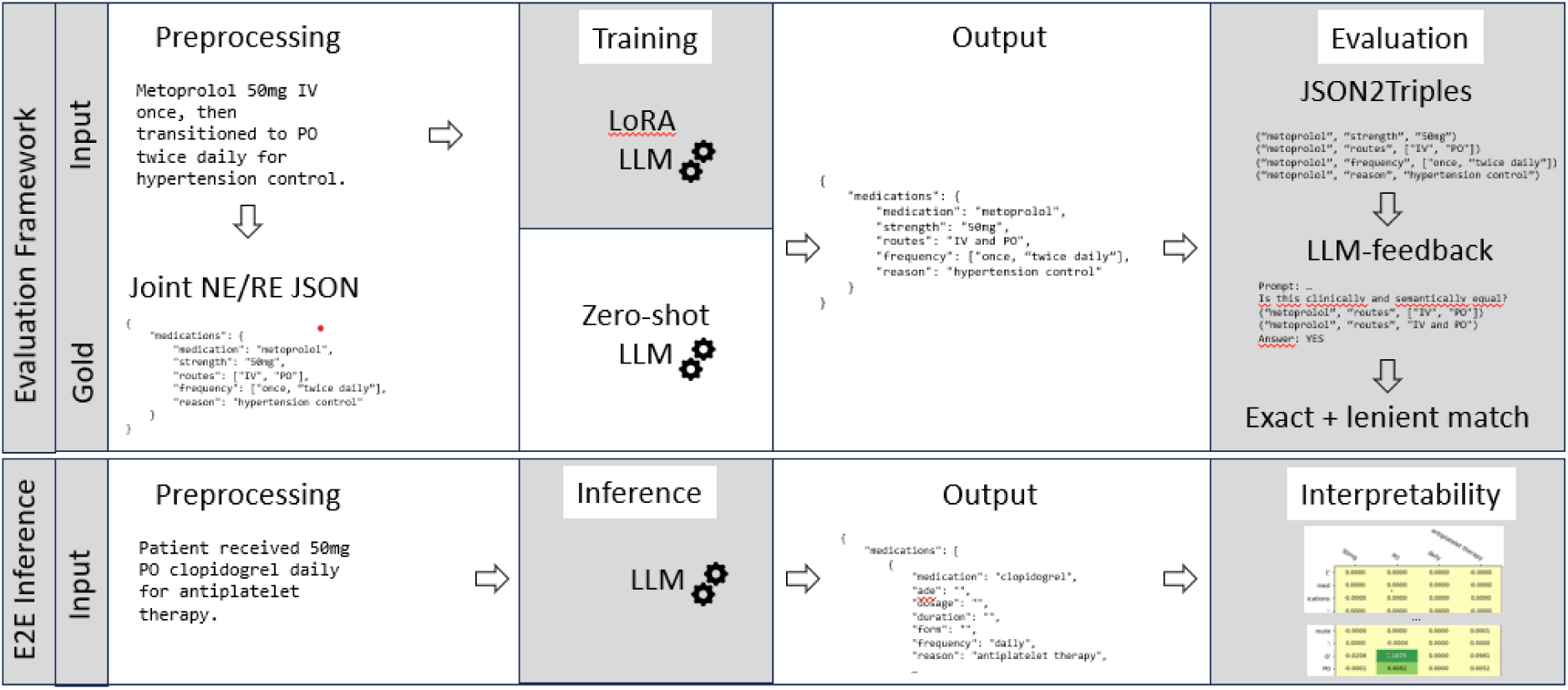
Graphical abstract: visualizing the evaluation framework and the e2e inference step.

### Data

All experiments were conducted on publicly available, manually annotated clinical routine corpora in English (n2c2 2018)[37] and German (CARDIO:DE)[47] language.

#### n2c2 2018 (track 2)

The corpus was distributed for the 2018 n2c2 shared task on adverse drug events and medication extraction in electronic health records and contains 505 English doctor’s letters from the MIMIC-III (Medical Information Mart for Intensive Care-III) clinical care database.[38] It was manually annotated with nine medication entity classes (*drug, strength, form, dosage, frequency, route, duration, reason, adverse drug event*) and eight medication relation classes (*drug-strength, drug-form, drug-dosage, drug-frequency, drug-route, drug-duration, drug-reason, drug-adverse drug event*).[37] The corpus is split into 303 letters in the training set and 202 letters in the test set. Overall, the corpus contains 83,869 entities and 59,810 relation annotations (cf. Suppl. Table 1, Suppl. Table 2).

#### CARDIO:DE v1.1

CARDIO:DE v 1.1 contains 500 German doctor’s letters from the cardiology department at the Heidelberg University Hospital.[47] The corpus was manually annotated with nine medication entity classes (*active ingredient, drug, strength, form, dosage, frequency, route, duration, reason*) and seven medication relation classes (*medication-strength, medication-form, medication-dosage, medication-frequency, medication-route, medication-duration, medication-reason*). The corpus is split into 400 letters in the training set and 100 letters in the test set. Overall CARDIO:DE contains 27,155 entities and 19,336 relation annotations (cf. Suppl. Table 3, Suppl. Table 4).

### Data preprocessing

Following best-practice in Dagdelen et al., we converted all annotations of the English and German datasets into uniform JSON strings.[34] The datasets contain a single doctor’s letter per file. We split each letter by newline (“\n”). We avoided sentence splitting, as manual evaluations showed frequent splitting errors for clinical texts. A JSON string with a single key “medications” and an empty list as a value is created. For each medication in the input text a JSON object with the medication name (entity) and all related information classes and their assigned entities is created and appended to the “medications” list (a comprehensive example, cf. Figure 3).

**Figure 3.**
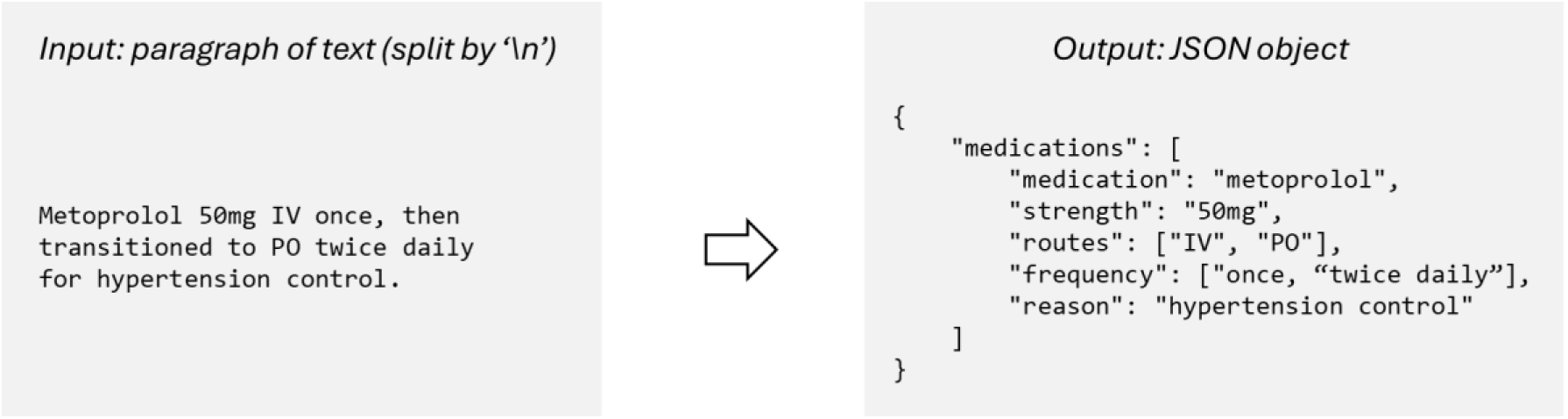
Data pre-processing: Converting a text snippet of a doctor’s letter into a JSON string. A JSON object with a medication (entity) Metoprolol and the related information classes (e.g. strength) and their assigned entities (e.g. 50mg)

However, related information of a medication name can appear across newline borders. Thus, we merged newline samples, if related medication information is contained in a neighboring sample (Table 1).

**Table 1.** Example of a merged sample. Strength and frequency information of Metoprolol is contained in a neighboring sample.

| Newline split samples | Merged sample | JSON |
| --- | --- | --- |
| Metoprolol to control<br>hypertension\n<br>50 mg twice daily. | Metoprolol to control<br>hypertension 50 mg twice<br>daily. | {<br>"medications": [<br>{<br>"medication": "metoprolol",<br>"strength": "50mg",<br>"routes": "",<br>"frequency": "twice daily",<br>"reason": "hypertension "<br>}<br>]<br>} |

This results in a data set which contains duplicate medications and medication relations. For the English n2c2 data set we count approximately 2.6% more relation information instances.

We also experimented with character offset information in the generated JSON object to define word boundaries and annotation boundaries, similar to traditional annotation schemes (XML, BRAT, etc.). However, manual evaluations showed that our LLMs did not reliably count correct character offsets for each medication information. To overcome this problem, for each medication in the input text we appended a medication object in the same textual order as in the text to the JSON string. However, due to the lack of character offset information, we could not use the official evaluation scripts of the n2c2 shared task, as they require this information.

Therefore, due to duplicates in the data set and missing character offset information, our results are not exactly comparable to the shared task baselines. Despite these drawbacks, following similar recent publications, the JSON representation allowed us to represent the complex information stored in e2e (concepts+ relations) annotations in a relatively compact format. The JSON strings can store multiple medications, with multiple related medication information. Furthermore, each relation information can be a list of strings containing multiple information values (e.g., “*’strength’: [‘5mg’, ‘10mg’]*”).

### Metrics

For all experiments we adopted the primary evaluation metric of the n2c2 shared task, lenient micro average F1-score per model and lenient F1-score per relation class. Lenient F1-score is 100% for relation class X if the drug name and the relation value is exactly matching or if there is an overlap between the gold relation value and the predicted relation value (lenient match examples, cf. Table 2). Furthermore, we present exact F1-score results in Suppl. Table 5 and Suppl. Table 6.

**Table 2.** Example gold standard samples and predictions of relation values. All predicted values are considered lenient match but not exact match.

| Gold value | Predicted value |
| --- | --- |
| *qhs* | *qhs (once a day at bed time)* |
| *25 mg / 3 ml* | *25 mg/3 ml* |
| *intraperitoneal bleeding* | *intraperitoneal bleeding from gist tumors* |
| *left eye* | *to left eye* |

### Local large language models

We used the official *Llama 3.1 base* model with 8 and 70 billion parameters (*meta-llama/Meta-Llama-3.1-[8B|70b]*) published by Meta Platforms for fine-tuning experiments, the respective instruct models (*meta-llama/Llama-3.1-[8B|70b]-Instruct*) for zero-shot experiments and *OpenBioLLM* with 8 billion parameters (*aaditya/Llama3-OpenBioLLM-8b*), a *meta-llama/Llama-3.1-8B-Instruct* model further-pretrained on biomedical texts. At the time of experiments, OpenBioLLM was the SOTA of medical domain-adapted LLMs (all used models, cf. Table 3).[54]

**Table 3.** Local large language models used for MIE experiments.

|  | Zero-shot | Fine-tuned (FT) |
| --- | --- | --- |
| Model | *Llama 3.1 instruct 8b/70b (4 bit)* | *Llama 3.1 base 8b/70b (4 bit)* |
|  | *OpenBioLLM 8b* | *OpenBioLLM 8b* |
| Training set | No training | Full training set |
| Training method | - | LoRA (r=16) |

For LoRA and QLoRA fine-tuning, we used the FastLanguageModel.get_peft_model implementation of the *Unsloth* library (cf. Low-rank adaptation (LoRA) and quantization).[55] In this work we utilize (i) QLoRA for training Llama 70b and (ii) LoRA to train all 8b LLMs.

For fine-tuning we used the complete training data set of n2c2 for the English experiments and CARDIO:DE for the German experiments (cf. Data).

#### Low-rank adaptation (LoRA) and quantized LoRA (QLoRA)

Due to the restricted computational resources available in a clinical environment and strict data protection regulations, it is necessary to employ efficient methods for training and inference of LLMs which possess billions of parameters.

LoRA is a PEFT method which freezes the LLM’s weights and injects trainable rank decomposition matrices into each transformer layer. This reduces the number of trainable parameters for fine-tuning and enables efficient task-switching by requiring only small, task-specific parameter updates. This makes LoRA a scalable solution for deploying large models in resource-constrained environments, such as the clinical domain (further details, cf. Hu et al, 2021) [8].

QLoRA further reduces memory usage during fine-tuning and extends LoRA by quantizing precision of the weight parameters of a LLM to 4-bit precision (for further information, cf. Dettmers et al, 2023) [11].

#### System prompt & structured output

We used a system prompt with instructions for the one-step e2e medication information extraction task. The model is prompted to extract all drug names including all related medication information in the order they appear in the paragraph. If >1 medications with the same name appear in the paragraph, we added a counter to the medication name in the order they appear in the input (system prompt cf. Figure 4).

**Figure 4.**
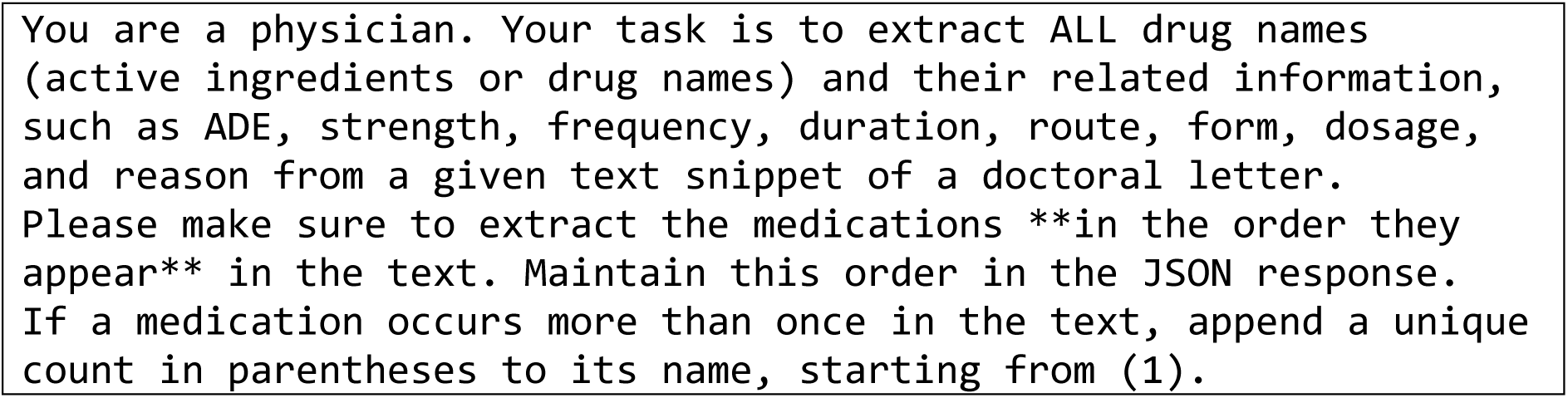
System prompt: Prompt used for all experiments (all LLMs, all datasets) except the ADE information, which is not available in the DE dataset.

Following current SOTA approaches, to steer LLMs to structured output, we added a pattern definition for the JSON output.[34] We used well-defined Pydantic object definitions as a format-restricting instruction in the systems prompt (https://pydantic.dev/articles/llm-intro). Using Pydantic classes simplifies structure definition and maintenance. Furthermore, we were able to add demonstrations in natural language for each medication information class (Figure 5**Fehler****! Verweisquelle konnte nicht gefunden werden.**).

**Figure 5.**
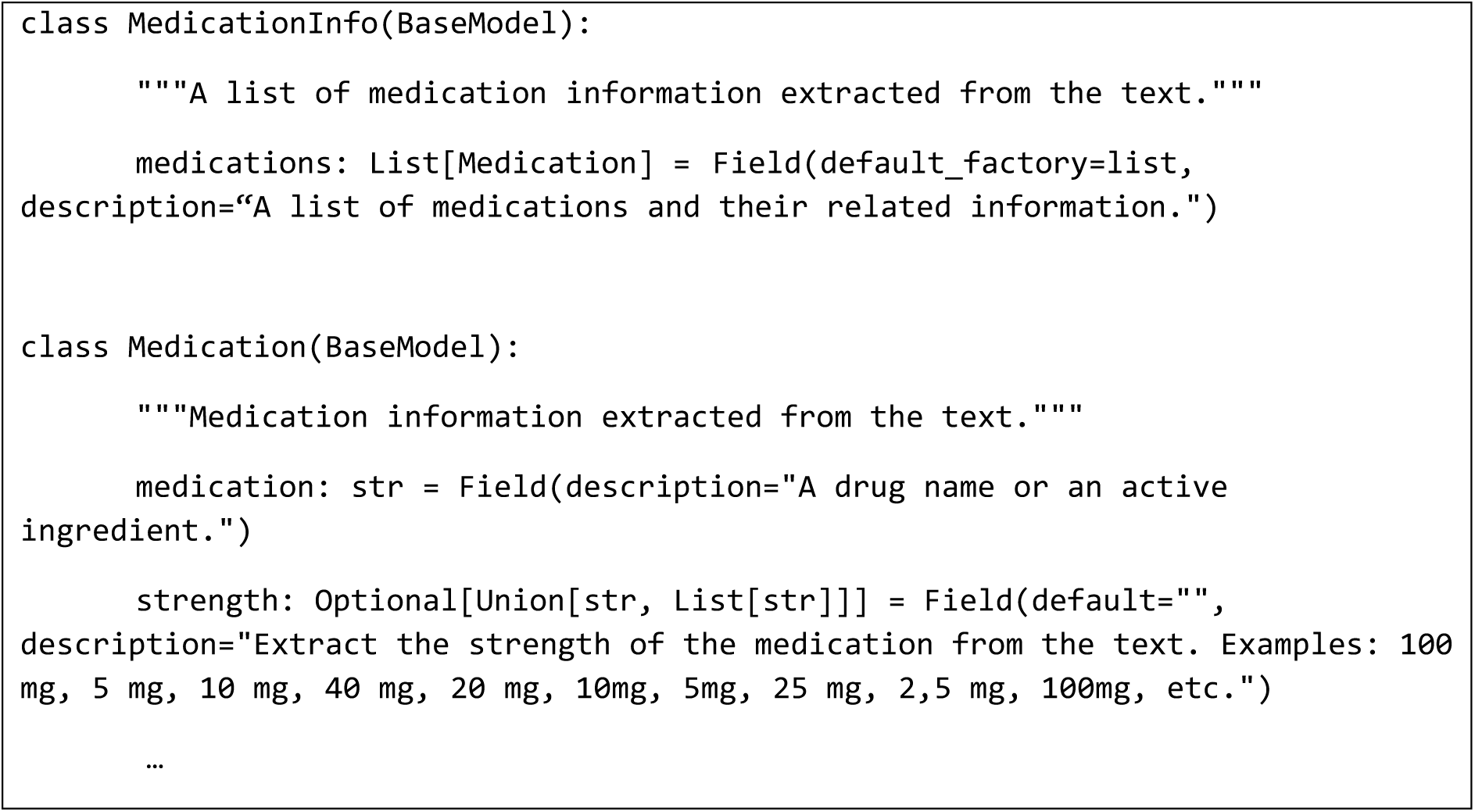
Format-restricting instruction: Pydantic objects to define two classes, the MedicationInfo class containing a list of Medication instances including medication name and related medication information (here only strength).

### Evaluation and feedback LLM

For a thorough evaluation we compared the prediction of each medication information class assigned to a medication in the JSON output with the gold standard annotation. For easier processing, we consistently converted each JSON of the gold standard and the prediction into a set of triplets, containing the medication class value (triplet head), the relation class (triplet predicate/class) and the relation value (triplet tail). Based on this structure we calculated precision, recall and F1-score per model (micro average) and per relation information class (cf. Table 4).

**Table 4.**
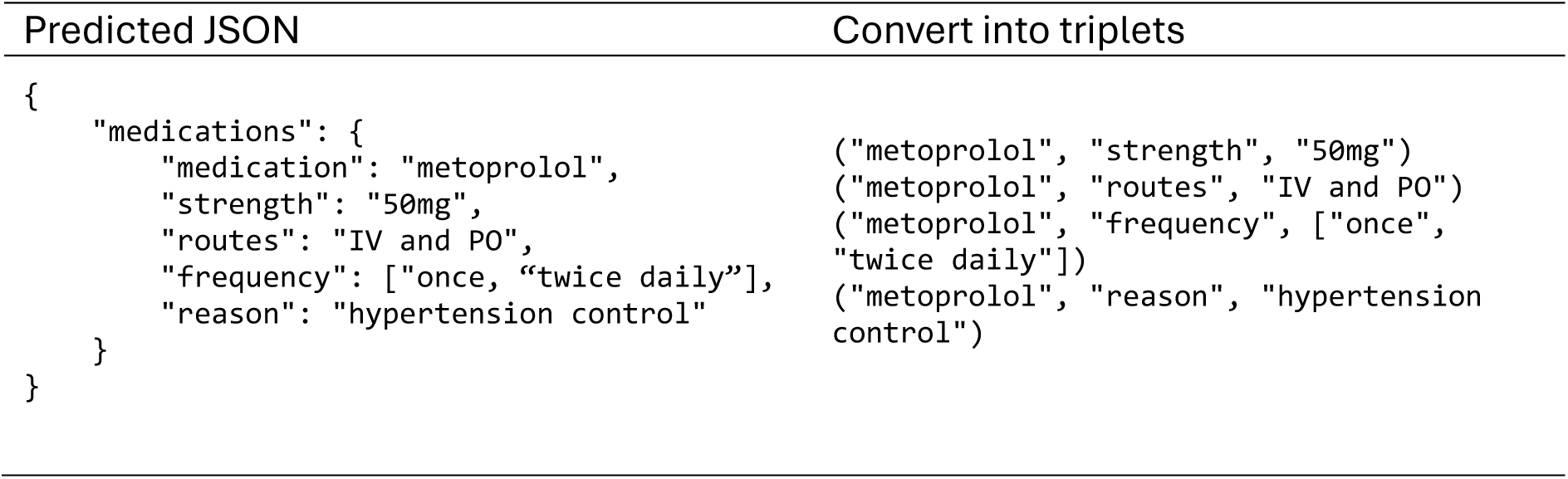
Converting predicted JSON strings into triplets for evaluation.

In contrast to traditional classification models, LLMs generate a sequence of tokens (in our setup: JSON strings). During manual evaluation using exact and lenient F1-scores, we discovered several patterns of false negative or false positive instances during manual analysis. For example, the model predicts (“metoprolol”, “routes”, “IV and PO”) and the gold standard is (“metoprolol”, “routes”, [“IV”, “PO”]). The gold standard defines the medication route as a list of two route strings, while the LLM predicted a simple string containing both route values connected by a coordinating conjunction “*and*”. However, the predicted value and the gold standard are clinically similar. Classifying such a case as false positive or false negatives would reflect mainly a technical issue, with little relevance for clinical routine. While handling such a simple case with heuristics and pattern matching seams feasible, we identified numerous patterns of false positives and negatives showcasing instances where a simple pattern matching would fail (cf. Table 5).

**Table 5.**
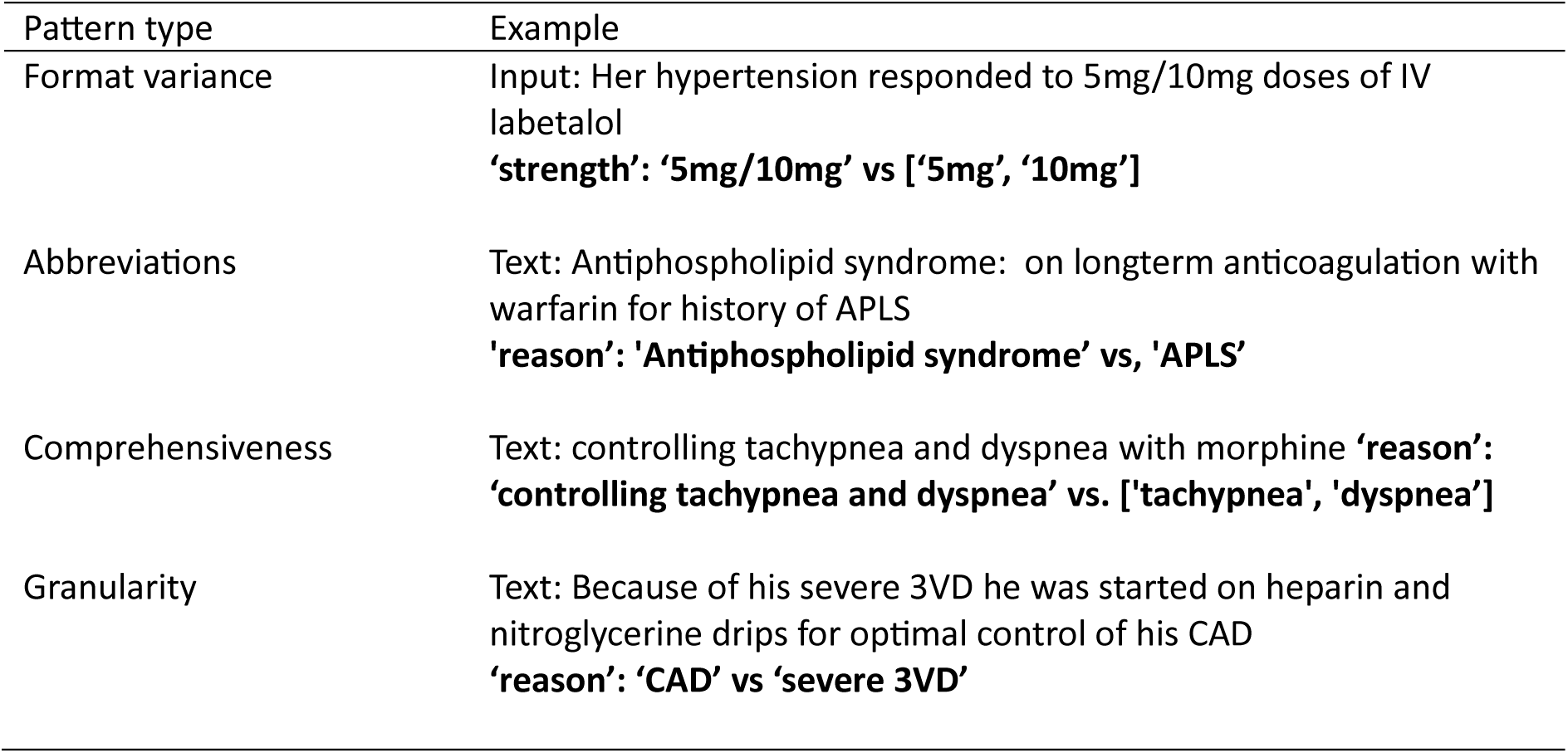
Error patterns of false positives and false negatives. First column: Pattern type, second column: pattern example, including the input paragraph and in bold face the relation class and the relation value gold vs predicted.

Because manual re-evaluation to identify these patterns is tedious and time-consuming, we followed recent research results using feedback LLMs for evaluation [10]. We employed the 4bit quantized Mistral-Large 123b (*unsloth/Mistral-Large-Instruct-2407-bnb-4bit*) as an expert LLM, to ensure diversity from our main MIE model. We performed a binary classification task: Given the input paragraph, the gold and the predicted value of false positives and false negatives, we prompted the model to classify the sample as *clinically similar/not similar* (system prompt cf. Suppl. Table 9). Cases of unrecognized or hallucinated medications or relation values are treated as valid false negatives or positives (Figure 6). We applied this approach for our best performing model (Llama 70b FT) for the English and German data set. A clinical expert manually reviewed the classification from a medical perspective. (Further details, cf. Suppl. section: Manual evaluation of the feedback LLM).

**Figure 6.**
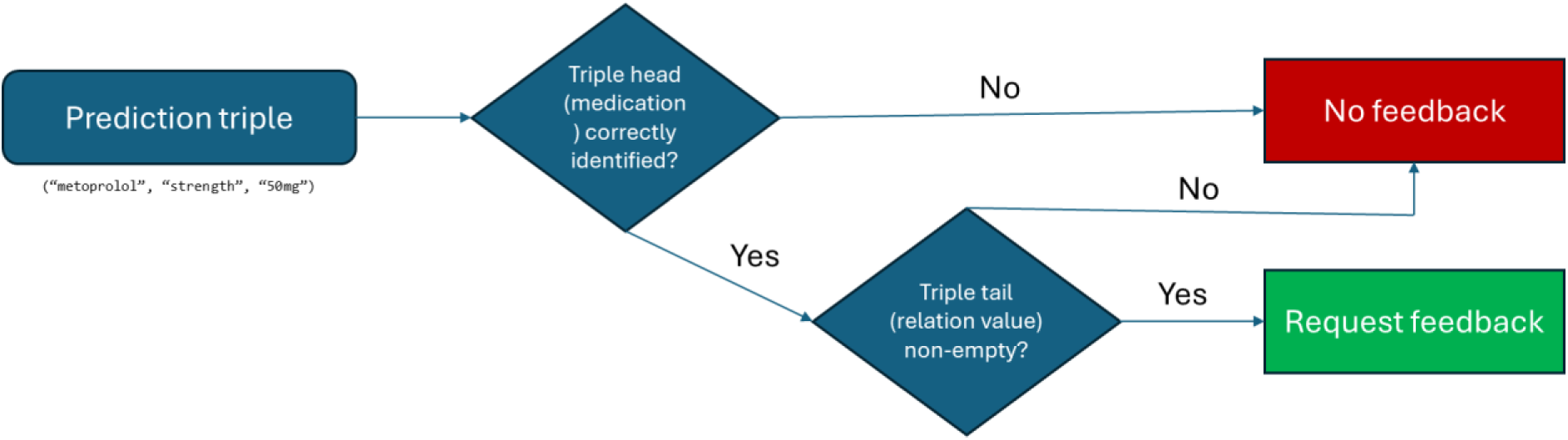
Feedback LLM: Pipeline to re-evaluate lenient false positive or false negatives predictions of the best performing model (Llama 70b) for English and German data.

### Shapley values

To increase trust in model predictions in the clinical domain, it is crucial to (1) understand the inner workings of a model (faithfulness) and to (2) evaluate how convincing a model interpretation is for a human observer (plausibility) [51]. This can increase trust in model predictions and support their application in the clinical routine.

In NLP, Shapley values became a valuable method for local model interpretation using saliency features.[52] They systematically quantify the influence of individual features (tokens or sequences of tokens) on model predictions. In our work, we utilize Shapley values in two ways: (1) to increase transparency for clinical users, and (2) from an engineering perspective, to identify errors in relation information extraction or false positive predictions. Derived from cooperative game theory, Shapley values determine input feature importance by averaging their marginal contributions across all possible feature combinations in predicting an output.[15] The Shapley value for a feature *i* in a model prediction is given by the equation:

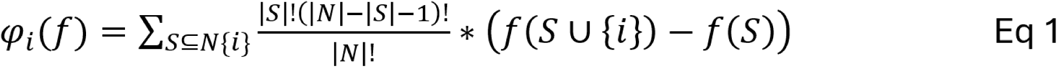

Where *f* is the prediction function, *S* is a subset of all features without feature *i*, and *N* is the set of all features.

In our work we use interpretability functionalities that are specifically designed to analyze the behavior of generative LLMs introduced in *Captum v 0.7*. This implementation offers various perturbation-based attribution methods and saliency map visualizations, including Shapley values [16]. The method generates Shapley values of each input token to each generated output token. Finally we provide further insights in model performance by presenting two use-cases using Shapley values for model interpretability.

## Results

In this section we present the one-step e2e joint NER and RE task results. We first present the LLM performance of zero-shot and SOTA experiments. Next, we present performance of fine-tuning experiments and further refine our results using feedback LLMs.

### Baselines

For the English and German data we conducted zero-shot experiments as a lower-bound baseline. Furthermore, for the English data we compare to two best-performing SOTA results for the MIE task, as reported by Modi et al, on the n2c2 data set, as our baseline. For the German CARDIO:DE data set, no MIE SOTA results had been reported so far. We thus conducted experiments using a vanilla transformer-based method, using a German domain-adapted BERT model (medBERT.de[56]).

### Zero-shot

#### n2c2

On n2c2, Llama 8b achieved the worst overall micro average F1-score with 62% (Table 6). Llama 70b increased performance by 15 percentage points. Both models showed worst performance for the complex classes *adverse drug event* (ADE) (27%;37%) and *reason* (29%;53%). However, for classes *dosage, frequency, route* and *strength,* F1-score was above 82%. Best-performing classes for the Llama 8b model were *frequency* (83%) and *route* (80%).

**Table 6.** Lenient F1-scores for n2c2 corpus for e2e MIE task. Comparing two SOTA baselines with zero-shot (zero) and FT (fine-tuned) Llama 8b and 70b and optimized evaluation using feedback LLM (FT feedback) for Llama 70b.

| Medication<br>information | Baseline |  | Llama 8b |  | Llama 70b |  |  |
| --- | --- | --- | --- | --- | --- | --- | --- |
|  | Wie et al. | El-Allaly et al. | zero | FT | zero | FT | FT (feedback) |
| **ade** | 48 | 65 | 27 | 70 | 37 | 73 | **75** |
| **dosage** | **94** | **94** | 68 | 93 | 82 | 93 | 93 |
| **duration** | 78 | 82 | 51 | 82 | 55 | 86 | **86** |
| **form** | 95 | 95 | 57 | 94 | 65 | 94 | **96** |
| **frequency** | **96** | 95 | 83 | 95 | 87 | 95 | **96** |
| **reason** | 58 | 78 | 29 | 79 | 53 | 82 | **84** |
| **route** | **94** | **94** | 80 | 94 | 87 | 93 | **94** |
| **strength** | **97** | 96 | 78 | 96 | 88 | 96 | **97** |
| **Micro avg.** | **89** | 91 | 62 | 91 | 77 | 92 | **93** |

#### CARDIO:DE

The micro average F1-score of Llama 8b on CARDIO:DE was only 5%, achieving a maximum of 14% for *frequency* and *strength (*Table 7). While micro average recall was 68%, precision only achieved 3%. A closer analysis confirmed this observation, demonstrating that Llama 8b exhibits substantial hallucinations, attributing medication information to nearly all input samples. In contrast, Llama 70b achieved a micro average F1-score of 71%. The mean performance was worse than for the English data set. However, *frequency* (87%) and *strength* (81%) achieved best F1-scores.

**Table 7.**
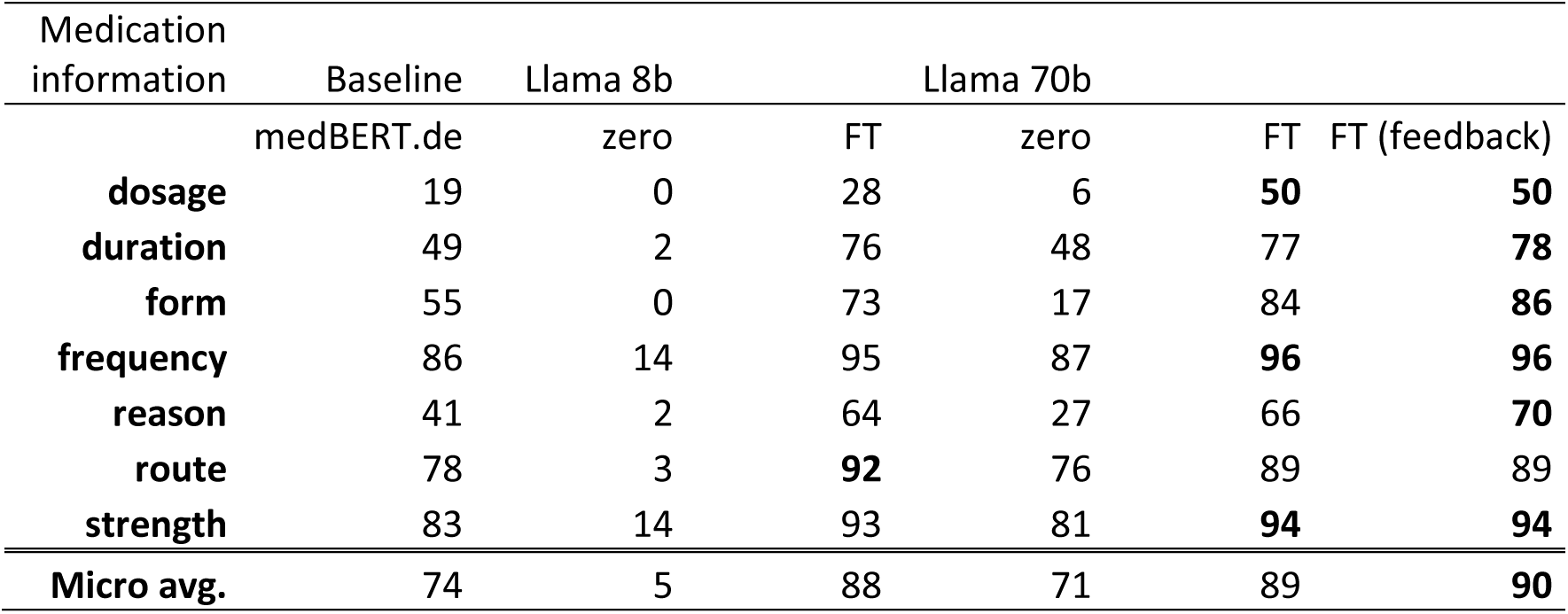
Lenient F1-scores for results CARDIO:DE corpus for e2e MIE task. Comparing SOTA baseline with zero-shot (zero) and FT (fine-tuned) Llama 8b and 70b and optimized evaluation using feedback LLM (FT feedback) for Llama 70b.

For Llama 8b and 70b the amount of malformed JSON outputs for either data set was marginal (0.002-0.2%). We were unable to reliably evaluate OpenBioLLM results, as the model frequently generated malformed JSON strings on the English and German dataset making automatic evaluation within our framework infeasible due to its dependence on well-structured JSON output.

### SOTA

#### n2c2

Wei et al. submitted the best performing system combining an RNN and CNN architecture in the n2c2 2018 shared task, with a micro average F1-score of 89%. Except *ADE*, *duration* and *reason* all class-wise F1-scores are >94%. For *ADE* the system achieved 48%; for *reason* 58%. This is only five percentage points above our Llama 70b in a zero-shot setting. Using SciBERT, El-Allaly et al. outperformed the system with 91% micro average F1-score. They especially improved results of *ADE* (65%), *duration* (82%) and *reason* (78%).

#### CARDIO:DE

We developed a pipeline including NER and RE using medBERT.de to (1) classify all medication entities and (2) all relations between the extracted entities. We achieved a micro average F1-score of 74%. This is only three percentage points above the Llama 70b zero-shot model. Best performing classes are *frequency* (86%), *route* (78%) and *strength* (83%). All other classes remained below 55%. The *dosage* class performed by far the weakest with 19% F1-score. Further analysis revealed that, unlike the English dataset, the German dataset typically includes *dosage* information within *frequency* information (e.g., *’1 tablet in the morning’* as *’1-0-0’*). Furthermore, with only 143 instances and a relatively low IAA of 73%, this class remains difficult to extract across all methods.

### Fine-tuned

#### n2c2

Fine-tuning Llama 8b on the n2c2 training set, the model achieved a micro average F1-score of 91%, outperforming the zero-shot baseline by 29% and on-par result with the baseline of El-Allaly et al (Table 6). However, Llama 8b outperforms this baseline for *ADE* (+5%) and *reason* (+1%). Llama 70b outperforms the SOTA with 92% micro-average F1-score. Particularly, for the complex classes *ADE* (+8%) and *reason* (+4%) and the *duration* class (+4%). For the classes *dosage, form* and *route* Llama 70b performs -1% worse than SOTA.

OpenBioLLM 8b did not improve results of Llama 8b (Suppl. Table 7). In contrast to the zero-shot experiments, all models, including OpenBioLLM, observed to the JSON format.

#### CARDIO:DE

Fine-tuning Llama 8b on the CARDIO:DE training set, the model achieved a micro average F1-score of 88%, substantially improving the results of the zero-shot model (+83%) and outperforming the BERT baseline by four percentage points (Table 7). Particularly for the complex *reason* class (+23%). Llama 70b further increased micro average F1-score to 89% and *reason* to 66%.

OpenBioLLM 8b achieved a micro average F1-score of just 7%, showing severe hallucinations, hence not benefiting from fine-tuning (Suppl. Table 8). However, similar to the English data, all models observed the JSON format.

### Using feedback pipeline

According to manual analysis of domain experts of false predictions, supposedly false positives and false negatives, in fact, frequently contained arguably clinically correct results. To automate manual evaluation of such instances, we used an external expert model (Mistral-Large 123b) to perform a binary classification for these samples compared to the gold standard (semantically *similar/not similar*). We applied this approach for our best performing model (fine-tuned Llama 70b) for the English and German data set.

#### n2c2

By using a feedback LLM to support automatic evaluation, micro-average F1-score could be increased to 93% (Table 6). Filtering false positives and false negatives particularly increased results for complex classes *ADE* (+2%) and *reason* (+2%), but as well for *form* (+2%), *frequency* (+1%) and *strength* (+1%). Hence, Llama 70b established a new SOTA for complex classes *ADE* (75%) and *reason* (84%), and further for *duration* (86%) and *form* (96%).

#### CARDIO:DE

The overall result was slightly increased to micro average F1-score 90%. Similar to n2c2, the biggest impact we observed for the complex class *reason* (+4%) and *form* (+2%) (Table 7). Hence, re-evaluation established a new SOTA result for the German data set.

### Interpretability

Our results show that LLMs achieved new SOTA results for MIE. However, due to their black-box nature, interpreting results remains challenging, limiting their adoption in the clinical domain [14]. While interpretability methods such as Shapley values are well-studied to explain ML models for structured data sources, they are increasingly used to interpret deep learning models for NLP tasks (further details, cf. Shapley values) [1]. However, their usage for generative LLMs remains widely understudied [16]. We therefore identified two use cases to apply Shapley values to further analyze LLMs in the context of our generative MIE task: (1) assessing the contributions of input tokens to relation information output tokens, (2) uncovering implicit knowledge on relation information. Below, we present two representative examples for each use case (further examples, cf. Suppl. section: Interpretability).

To evaluate whether input token contributions align with the relation information output tokens, we investigated a key edge case in MIE. Since our MIE task, aside from JSON syntax, reproduces input tokens in the output, their contribution to each output token may appear trivial. For instance, if the input contains the relation information for strength “*5 mg*” and the model correctly predicts “*’strength’: ‘5 mg*’” in the JSON string, the contribution alignment seems straightforward. However, when the same strength relation value (cf. Figure 7) appears twice in the input, each linked to a different medication name, we can analyze whether the first occurrence correctly maps to the first medication, and the second to the corresponding second medication.

**Figure 7.**
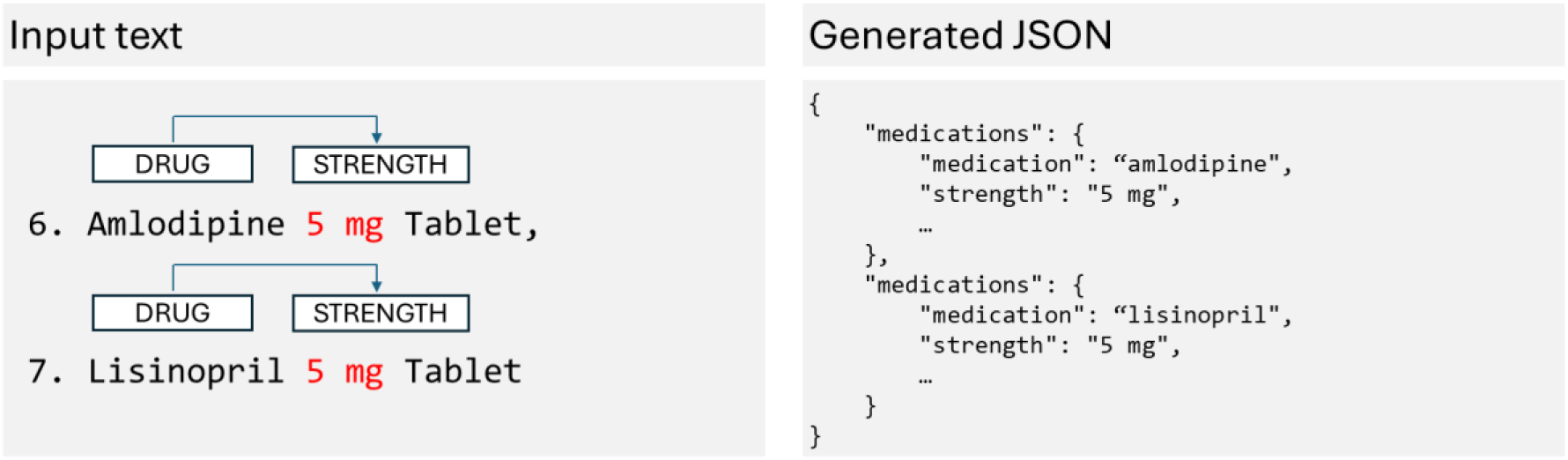
Use case 1 input data: (left) Example input text containing two medications (Amlodipine, Lisinopril) each related to a similar strength value (5 mg). (right) Corresponding generated JSON output snippet containing the medication names and the strength values.

Investigating the contributions of input tokens, as indicated via Shapley values (Figure 8) for the Llama 70b FT model, we observed that the first occurrence of the strength token correctly contributes to the strength value of the first medication (Amlodipine) in the JSON output. Specifically, the first “*5 mg*” token is linked to Amlodipine, showing the highest contribution to its JSON entry, while the second “*5 mg*” corresponds to Lisinopril, with the highest contribution to its respective JSON entry. In both cases we observed that the strength input token has the strongest positive or negative contribution to the initializing quote of the respective strength value. The contribution of the respective digit is significantly lower.

**Figure 8.**
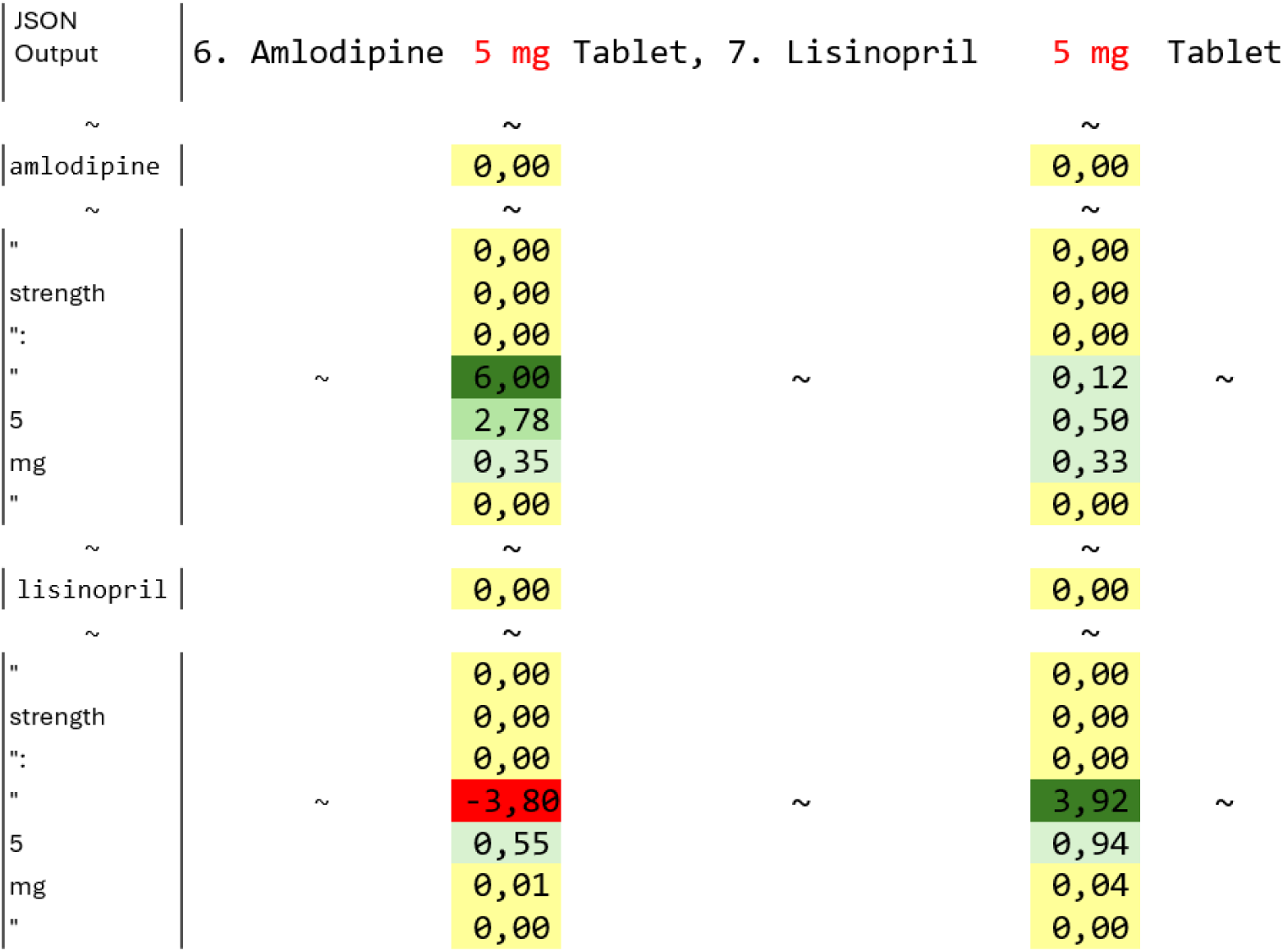
Use case 1 Shapley values: Visualizing approximate Shapley values for the strength token of the input text “6. Amlodipine 5 mg Tablet, 7. Lisinopril 5 mg Tablet” and the generated JSON token for the strength relation class and value. Complete output cf. Suppl. Figure 3

We manually evaluated various examples for this scenario. All of them indicated that the model correctly assigns related medication information to the correct medication instances, though differences in token contributions vary (cf. n Suppl. Figure 1 Suppl. Figure 3 - 9).

The second use case uncovers that LLMs possess implicit knowledge regarding relation information, even when they do not explicitly generate this information in the JSON output. We therefore selected a representative instance of the n2c2 dataset. This instance contains an *ADE* (*acute renal insufficiency*) which was not generated in the JSON output by our Llama 8b FT model (Figure 9).

**Figure 9.**
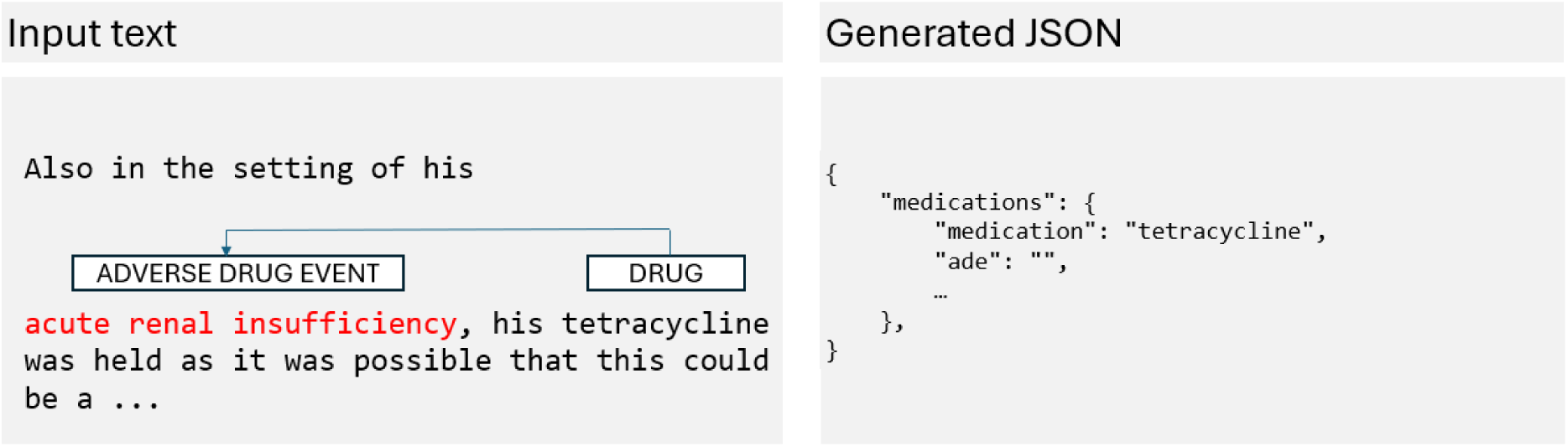
Use case 2 input data: (left) Example input text containing ADE of the medication Tetracycline. (right) Corresponding generated JSON output snippet containing the medication name and the empty ADE value.

We therefore calculated Shapley values only of the *ADE* tokens for this instance (Figure 10). These tokens contribute negatively to the empty ADE output of the model. Upon further analysis, we found that the output probability of the empty string decreased to 61%. This suggests that while the model correctly recognizes the input as an *ADE*, the probability assigned to the corresponding *ADE* relation in the output remains insufficient for its explicit generation. We conducted additional manual evaluations, which revealed a similar behavior across various relation classes, as shown in Suppl. Figure 10-19.

**Figure 10.**
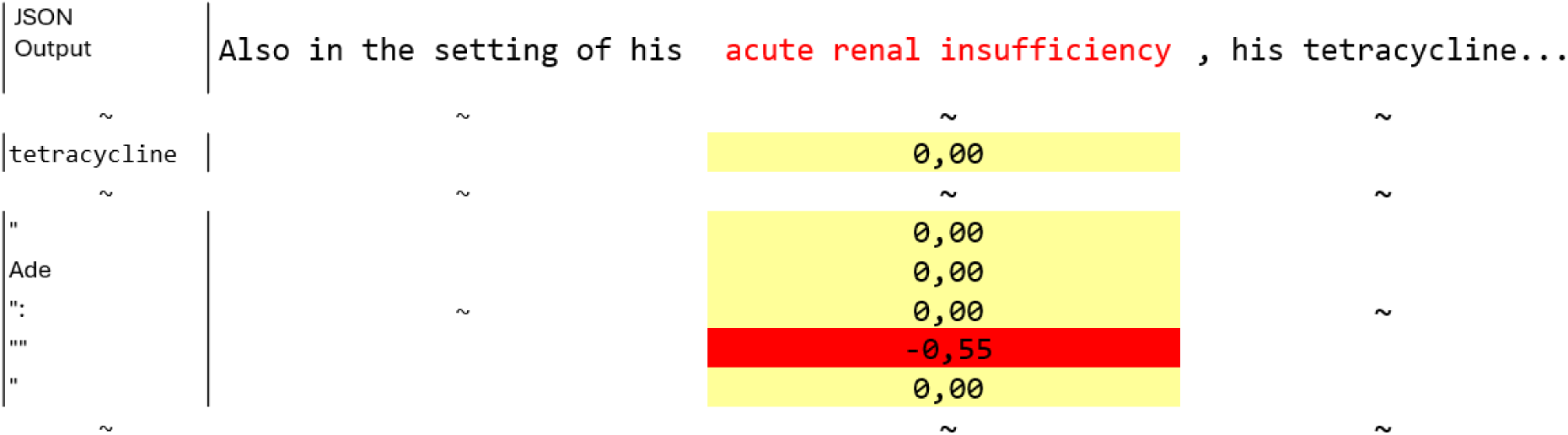
Use case 2 Shapley values: Visualizing approximate Shapley values for the ADE token of the input text “Also in the setting of his acute renal insufficiency, his tetracycline was held as it was possible that this could be a …” and the generated JSON token for the ADE relation class and value.

In a future study, we plan to use Shapley values to uncover implicit relation information in LLMs in a clinical scenario. Through a graphical user interface, physicians will be presented with instances where input tokens have strong negative contributions to empty relation information outputs. These instances will be provided for further analysis, helping to identify and investigate potential false negative predictions.

## Discussion & Conclusion

In this section, we discuss and conclude our empirical results in relation to the challenges and proposed solutions presented in the Introduction.

Ch 1. **Demand of clinical experts** Based on the observation that LLMs contain clinical knowledge, we evaluated both open-source foundation LLMs based on Llama and the domain-adapted biomedical OpenBioLLM, comparing their performance in zero-shot and fine-tuned setups, evaluated on well-curated English and German datasets.
**Finding:** In a zero-shot setup on English data, Llama 8b and 70b showed strong performance but clearly lagged behind SOTA. On German data, Llama 8b showed strong hallucinations, while Llama 70b performs more stably but is still inadequate for a clinical setup.
**Finding:** In a fine-tuning setup on English data, Llama models define a new SOTA for complex classes such as *ADE*, *reason*, and *duration*. On German data, both models show an overall lower performance in comparison to English data, however, they set a new benchmark for all classes on German data.
**Finding:** In a zero-shot setup OpenBioLLM frequently generated malformed structured format predictions in English and German. In a fine-tuning setup OpenBioLLM showed reliable structured output format and comparable performance to Llama on English data. However, while structured output was generated reliably on German data, the model consistently produced hallucinations.
**Conclusion**: Fine-tuning improves performance particularly for open-source foundation LLMs with Llama 70b achieving top performance, optimizing the return of invest by reducing the demand for clinical expertise for creating training data. In contrast, OpenBioLLM lacks robustness in structured output generation and frequently produces hallucinations.
Ch 2. **Limited and costly time resources** We defined our MIE task as a one-step e2e joint NER and RE task on two gold standard data sets with well-defined structured output formats and used a feedback LLM to support automatic evaluation.
**Finding:** In a zero-shot setup, both Llama models reliably produced structured output. In a fine-tuning setup, all models consistently followed the defined output structure.
**Finding**: The generative capabilities of LLMs simplified the e2e pipeline, significantly reducing both development and maintenance efforts. However, all LLMs occasionally deviate from the gold standard, while frequently remaining clinically correct. This motivated us to leverage feedback LLMs to assist in assessing critical predictions.
**Finding:** Feedback LLMs helped to speed-up the oftentimes complex evaluation of LLM outputs and further helped identify frequent false positives and negatives that were actually correct. Additionally, they supported our findings that LLMs surpass the current SOTA.
**Conclusion:** All Llama models adhered to the output structure using format-restricting instructions. OpenBioLLM needed fine-tuning for reliable structured output. While the generative nature of LLMs allowed us to build e2e MIE solutions requiring less development and maintenance effort, it also made evaluation more laborious. Thus, to support automatic evaluation, we present feedback LLMs to assist in in fine-grained output evaluation.
Ch 3. **Limited clinical IT resources** Leveraging PEFT fine-tuning based on LoRA and quantization, we used smaller open-source LLMs for our MIE task.
**Finding:** We could fine-tune a Llama 70b model using 4bit QLoRA on our clinical infrastructure using a single NVIDIA H100 (using 48GB of 96GB VRAM). 8b models could be fine-tuned using a single NVIDIA RTX6000 (using 14GB of 24GB VRAM).
**Finding**: Fine-tuning LLMs as clinical expert LLMs achieved new SOTA results for the 70b model, while the smallest 8b model performs only slightly worse. This further reduces IT infrastructure requirements, while generating reliable structured output.
**Conclusion:** All Llama models are resource-efficient to fine-tune with 8b models being highly IT-friendly. 8b models perform well overall, but 70b models are superior for complex classes, particularly on German data.
Ch 4. **Transparent and comprehensive predictions** We used Shapley values and saliency maps optimized for generative LLMs to support model evaluation and to support transparency of model predictions.
**Finding:** Similar to traditional applications, Shapley values remain useful for measuring token-level input contributions to generative LLM predictions, providing faithfulness at the token level. However, recent research indicates this token-level faithfulness does not necessarily reflect the model’s underlying reasoning process, but rather measures self-consistency – an essential property for clinical MIE. [57]
**Finding:** In our scenario Shapley values highlight input token contributions to extracted structured output predictions. For our two use cases this supports understanding the relation extraction performance of LLMs and allowed us to identify implicit clinical knowledge, in cases where the model makes false predictions.
**Conclusion:** Combined with output probabilities, Shapley values and saliency maps improve the prediction transparency of LLMs. Importantly, saliency maps can aid physicians in evaluating predictions, supporting informed clinical decision-making.

## Data availability

For the English MIE task, we used the n2c2 2018 (track 2) dataset. It was released as part of the 2018 National NLP Clinical Challenges (n2c2) shared task. Due to patient privacy regulations, access to the dataset requires approval through the n2c2 Data Use Agreement (DUA). Researchers can request access via the official n2c2 portal (https://n2c2.dbmi.hms.harvard.edu/2018-challenge).[37]

For the German MIE task, we used CARDIO:DE (v. 1.1), a distributable German corpus containing 500 cardiovascular doctor’s letters from the clinical routine, for all our experiments (available with a signed DUA: https://doi.org/10.11588/data/AFYQDY). Annotations of the held-out datasets are not publicly available as authors of CARDIO:DE use it for shared task competitions. But they are available from the corresponding author on reasonable request. For more details about the dataset, preprocessing steps, data annotation, and distribution, cf. Richter-Pechanski *et al*. [47].

## Data Availability

For the English MIE task, we used the n2c2 2018 (track 2) dataset. It was released as part of the 2018 National NLP Clinical Challenges (n2c2) shared task. Due to patient privacy regulations, access to the dataset requires approval through the n2c2 Data Use Agreement (DUA). Researchers can request access via the official n2c2 portal (https://n2c2.dbmi.hms.harvard.edu/2018-challenge).
For the German MIE task, we used CARDIO:DE (v. 1.1), a distributable German corpus containing 500 cardiovascular doctors letters from the clinical routine, for all our experiments (available with a signed DUA: https://doi.org/10.11588/data/AFYQDY). Annotations of the held-out datasets are not publicly available as authors of CARDIO:DE use it for shared task competitions. But they are available from the corresponding author on reasonable request. For more details about the dataset, preprocessing steps, data annotation, and distribution, cf. Richter-Pechanski et al.

https://doi.org/10.11588/data/AFYQDY

## Acknowledgments

The authors disclosed receipt of the following financial support for the research, authorship, and/or publication of this article: The work of PR-P, NG and CD was kindly supported by the DFG (German Research Foundation, DFG D1501/14-1) and Informatics for Life funded by the Klaus Tschira Foundation and for PR-P and CD by the BMBF-funded HiGHmed consortium (www.highmed.org, consortium of the Medical Informatics Initiative Germany).

We would like to thank all members of the Dieterich Lab and the Heidelberg Natural Language Processing Group for their great input and insightful discussions.

## Author contributions

**Phillip Richter-Pechanski:** Conceptualization, Methodology, Software, Validation, Formal analysis, Investigation, Resources, Data Curation, Writing - Original Draft, Visualization. **Marvin Seiferling:** Validation, Formal analysis. **Christina Kiriaku:** Validation, Resources, Data Curation. **Dominic Schwab:** Validation, Resources, Data Curation. **Nicolas Geis:** Resources, Data Curation, Funding acquisition, Writing - Review & Editing. **Christoph Dieterich:** Project administration, Supervision, Resources, Funding acquisition, Writing - Review & Editing. **Anette Frank:** Project administration, Supervision, Conceptualization, Methodology, Formal analysis, Investigation, Writing - Review & Editing.

## Ethics statement

The authors state that this study complies with the Declaration of Helsinki. Our task has been performed with respect to Section 46 Abs.2 Nr.2a (LKHG) and Section 13 Abs.1 Landesdatenschutzgesetz BW. In this context, we had the possibility to use the data for the purpose of optimizing internal clinical procedures.

## Supplementary material

**Suppl. Table 1.**
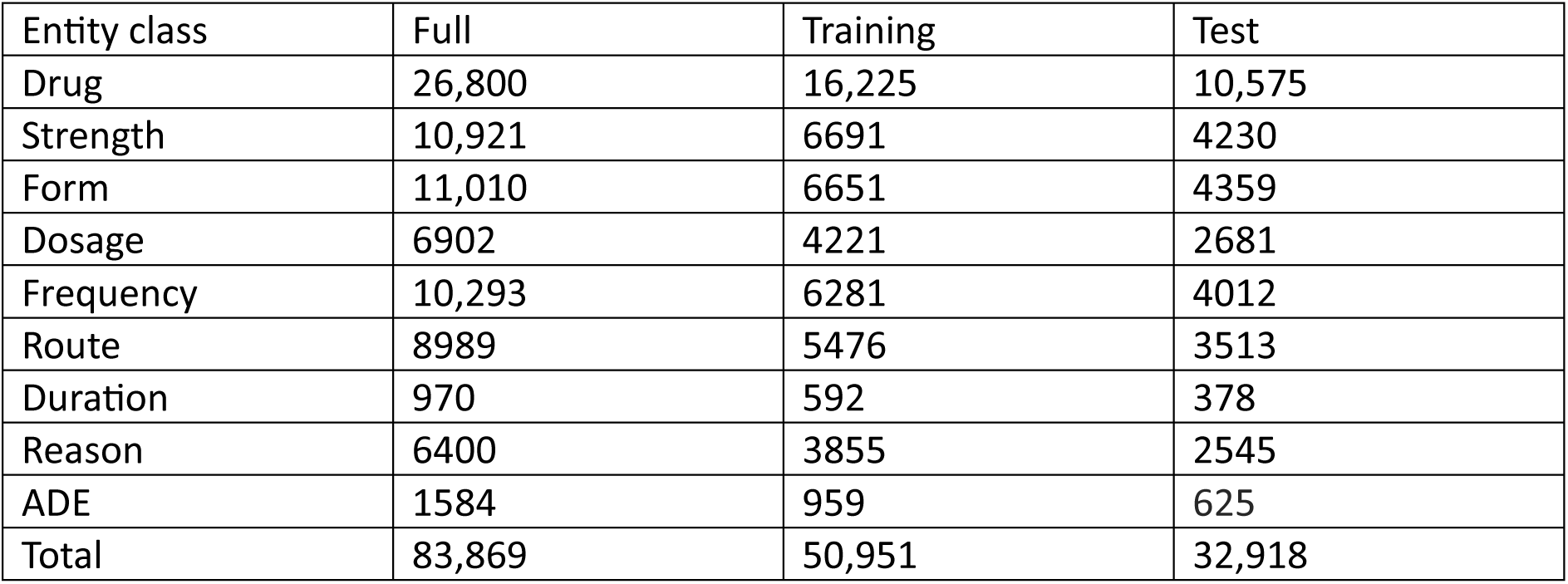
n2c2 2018 (track 2) corpus, annotated entity classes.

| Entity class | Full | Training | Test |
| --- | --- | --- | --- |
| Drug | 26,800 | 16,225 | 10,575 |
| Strength | 10,921 | 6691 | 4230 |
| Form | 11,010 | 6651 | 4359 |
| Dosage | 6902 | 4221 | 2681 |
| Frequency | 10,293 | 6281 | 4012 |
| Route | 8989 | 5476 | 3513 |
| Duration | 970 | 592 | 378 |
| Reason | 6400 | 3855 | 2545 |
| ADE | 1584 | 959 | 625 |
| Total | 83,869 | 50,951 | 32,918 |

**Suppl. Table 2.** n2c2 2018 (track 2) corpus, annotated relations.

| Relation class | Full | Training | Test |
| --- | --- | --- | --- |
| Drug-Strength | 10,946 | 6702 | 4244 |
| Drug-Form | 11,028 | 6654 | 4374 |
| Drug-Dosage | 6920 | 4225 | 2695 |
| Drug-Frequency | 10,344 | 6310 | 4034 |
| Drug-Route | 9084 | 5538 | 3546 |
| Drug-Duration | 1069 | 643 | 426 |
| Drug-Reason | 8579 | 5169 | 3410 |
| Drug-ADE | 1840 | 1107 | 733 |
| Total | 59,810 | 36,384 | 23,462 |

**Suppl. Table 3.** CARDIO:DE corpus, annotated entity classes.

| Entity class | Full | Training | Test |
| --- | --- | --- | --- |
| Active Ingredient (AI) | 7540 | 6032 | 1508 |
| Drug | 2195 | 1747 | 448 |
| Strength | 6442 | 5071 | 1371 |
| Form | 181 | 152 | 29 |
| Dosage | 143 | 127 | 16 |
| Frequency | 6501 | 5135 | 1366 |
| Route | 567 | 452 | 115 |
| Duration | 1596 | 1284 | 312 |
| Reason | 1990 | 1631 | 359 |
| Total | 27,155 | 21,631 | 5524 |

**Suppl. Table 4.** CARDIO:DE corpus, annotated relations.

| Relation class | Full | Training | Test |
| --- | --- | --- | --- |
| AI/Drug-Strength | 6510 | 5093 | 1417 |
| AI/Drug-Form | 185 | 155 | 30 |
| AI/Drug-Dosage | 146 | 130 | 16 |
| AI/Drug-Frequency | 6658 | 5231 | 1427 |
| AI/Drug-Route | 1385 | 1058 | 327 |
| AI/Drug-Duration | 1850 | 1487 | 363 |
| AI/Drug-Reason | 2602 | 2116 | 486 |
| Total | 19,336 | 15,270 | 4066 |

**Suppl. Table 5.** Exact F1-scores for n2c2 corpus for e2e MIE task. Zero-shot (zero) and FT (fine-tuned) Llama 8b and 70b.

| Medication<br>information | Llama 8b |  | Llama 70b |  |
| --- | --- | --- | --- | --- |
|  | zero | FT | zero | FT |
| **ade** | 22 | 63 | 29 | 67 |
| **dosage** | 24 | 90 | 45 | 90 |
| **duration** | 20 | 72 | 37 | 71 |
| **form** | 51 | 91 | 61 | 91 |
| **frequency** | 58 | 88 | 69 | 89 |
| **reason** | 21 | 69 | 42 | 73 |
| **route** | 79 | 93 | 86 | 92 |
| **strength** | 74 | 92 | 83 | 91 |
| **Micro avg.** | 49 | 86 | 65 | 87 |

**Suppl. Table 6.** Exact F1-scores for CARDIODE corpus for e2e MIE task. Zero-shot (zero) and FT (fine-tuned) Llama 8b and 70b.

| Medication<br>information | Llama 8b |  | Llama 70b |  |
| --- | --- | --- | --- | --- |
|  | zero | FT | zero | FT |
| **dosage** | 0 | 28 | 3 | 50 |
| **duration** | 1 | 59 | 33 | 61 |
| **form** | 0 | 73 | 17 | 84 |
| **frequency** | 14 | 93 | 84 | 94 |
| **reason** | 1 | 49 | 18 | 51 |
| **route** | 3 | 91 | 74 | 89 |
| **strength** | 14 | 92 | 80 | 93 |
| **Micro avg.** | 5 | 84 | 67 | 84 |

**Suppl. Table 7.** Lenient F1-scores for n2c2 corpus for e2e MIE task. Zero-shot (zero) and FT (fine-tuned) Openbiollm 8b.

| Medication<br>information | Openbiollm 8b |  |
| --- | --- | --- |
|  | zero | FT |
| **ade** | - | 69 |
| **dosage** | - | 92 |
| **duration** | - | 83 |
| **form** | - | 94 |
| **frequency** | - | 95 |
| **reason** | - | 78 |
| **route** | - | 93 |
| **strength** | - | 96 |
| **Micro avg.** | - | 91 |

**Suppl. Table 8.**
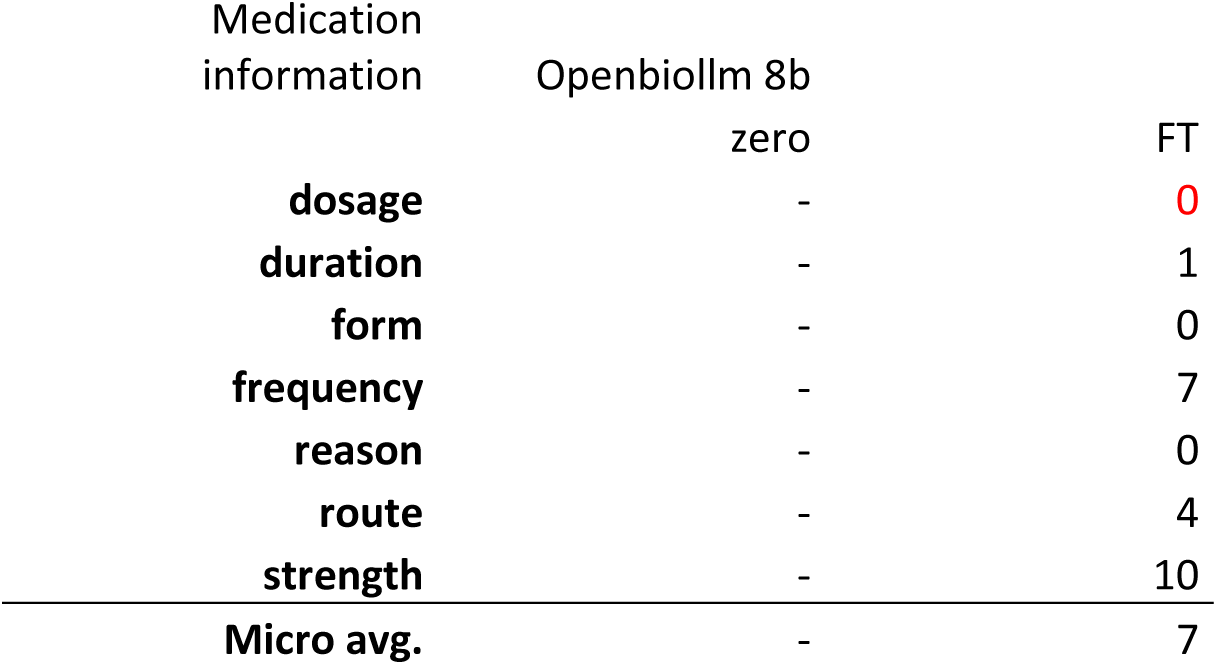
Lenient F1-scores for CARDIO:DE corpus for e2e MIE task. Zero-shot (zero) and FT (fine-tuned) Openbiollm 8b.

**Suppl. Table 9.**
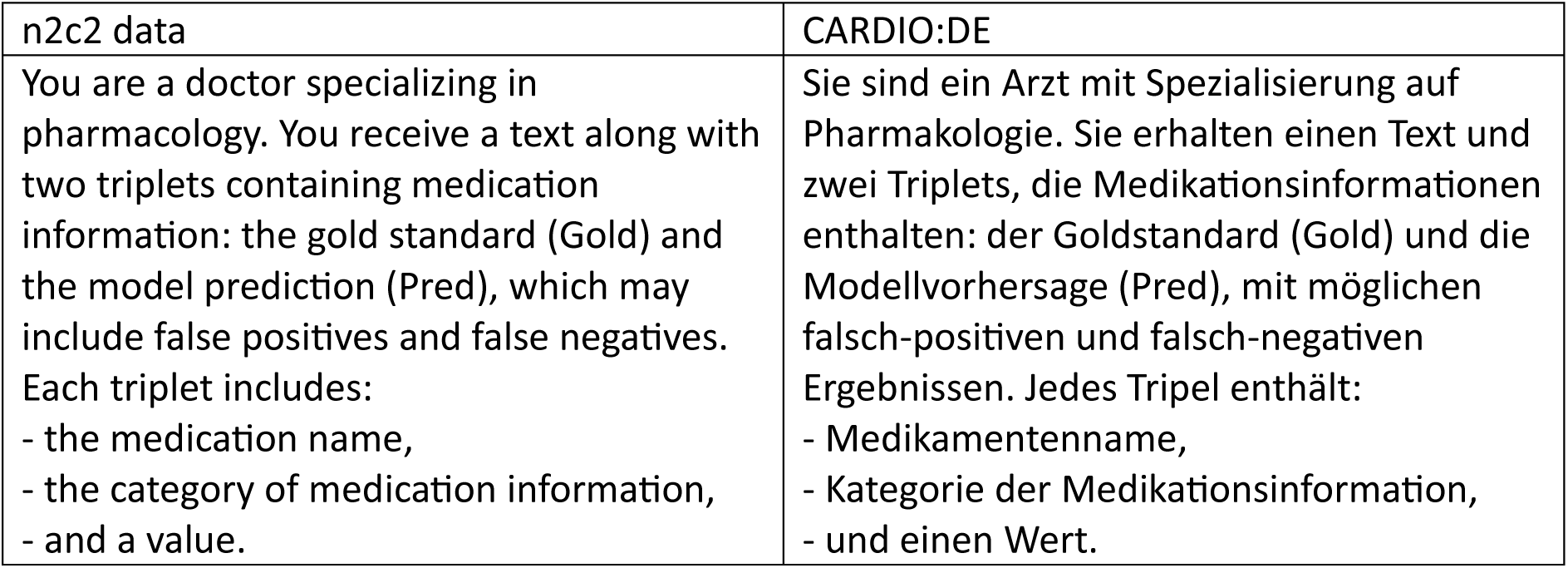
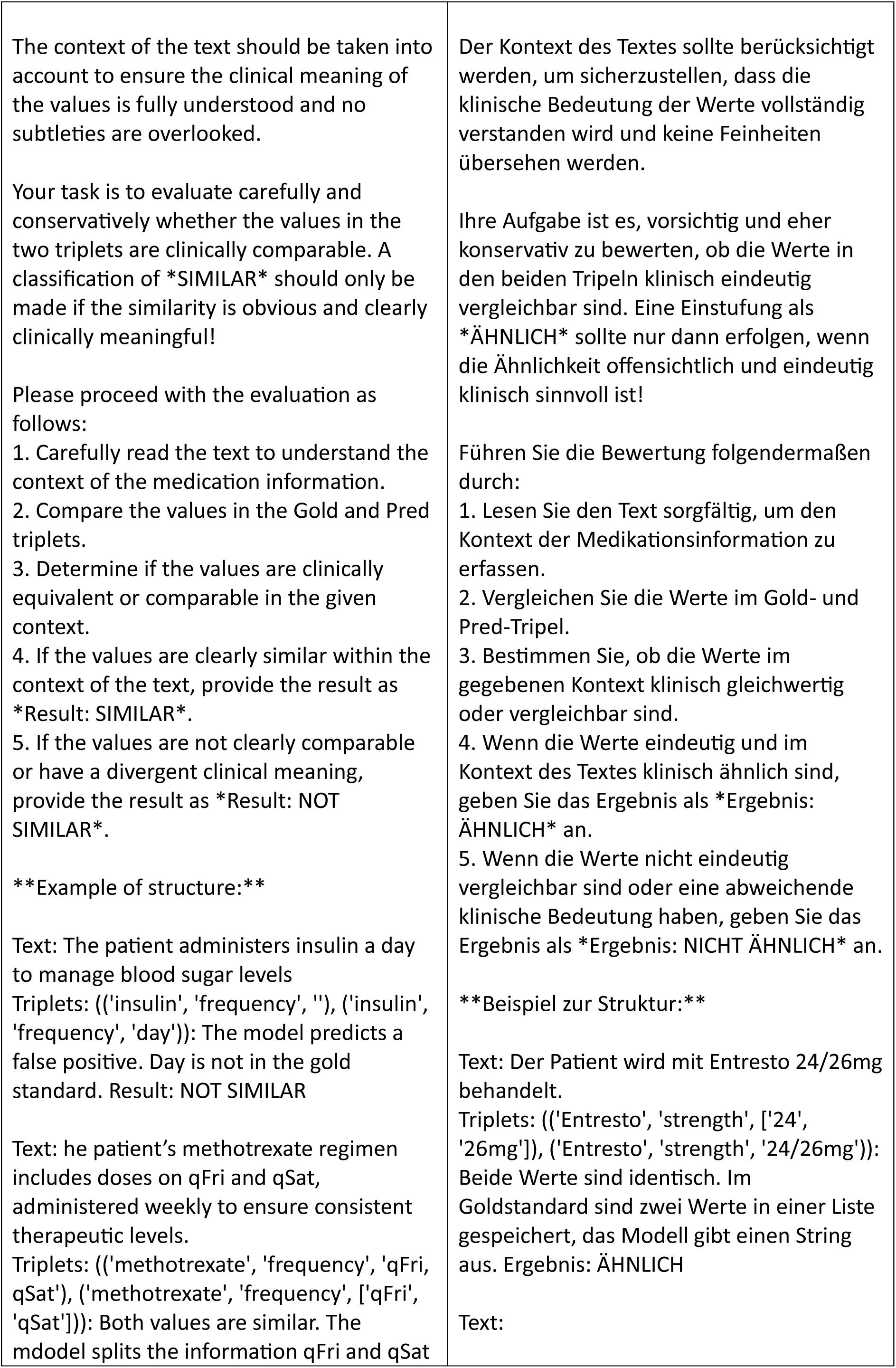
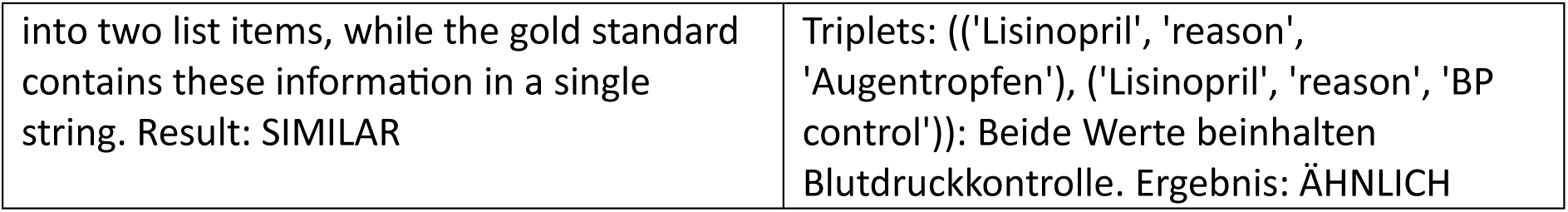
System prompt for the feedback LLM task for English and German data.

### Suppl. section: Manual evaluation of the feedback LLM

To evaluate the performance of the feedback LLM we conducted manual evaluation of a subset of all predicted instances on the EN dataset. For each relation class, and separately for false positive and false negative samples, if available, we selected five instances classified as SIMILAR and five instances classified as NOT SIMILAR by the feedback LLM (overall 149 instances). A clinical expert, presented with the gold standard, the LLM prediction and the feedback LLM classification, evaluated each instance to determine if it was classified correctly from a clinical perspective.

The clinical expert agreed with the feedback LLM’s “SIMILAR” classification in 94% of cases. In contrast, he only agreed with the feedback LLM’s “NOT SIMILAR” classification in 44% of cases. However, the expert often considered instances labeled as NOT SIMILAR to be SIMILAR as well. This suggests that the feedback model acts as a more cautious classifier, which can be advantageous in a clinical context, as the expert frequently reclassified ‘NOT SIMILAR’ instances as ‘SIMILAR’. It also highlights the strong performance of our medication information classifier.

### Suppl. section: Interpretability

In this section we present further examples for the application of Shapley values for use case: (1) assessing input token contribution to relation information output token (cf. Suppl. Figure 3 - 9), (2) uncovering implicit knowledge on relation information (cf. Suppl. Figure 10-18).

#### Use case 1

The following examples demonstrate the contributions of input tokens to relation information output tokens. To show the ability of LLMs if they correctly assign relation information to medications we investigate a specific edge case: the same relation value appears twice in the input, each linked to a different medication name. Thus, we can analyze whether the first occurrence correctly maps to the first medication, and the second to the corresponding second medication.

##### Example 1

**Suppl. Figure 1.**
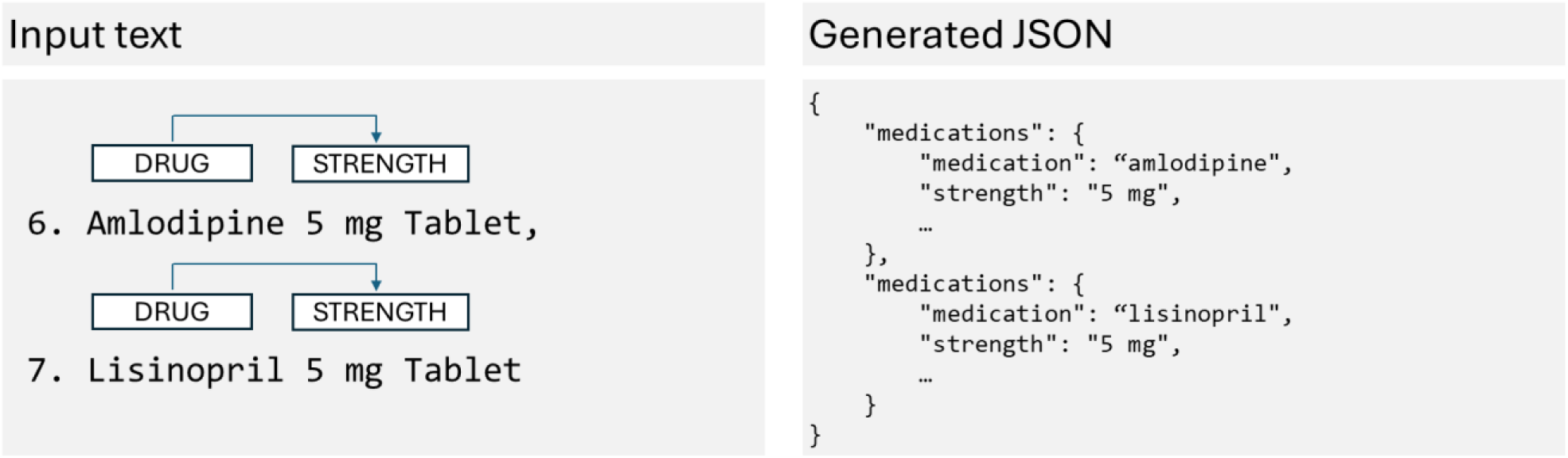
(left) Example input text containing two medications (Amlodipine, Lisinopril) each related to a similar strength value (5 mg). (right) Corresponding generated JSON output snippet containing the medication names and the strength values.

**Suppl. Figure 2.**
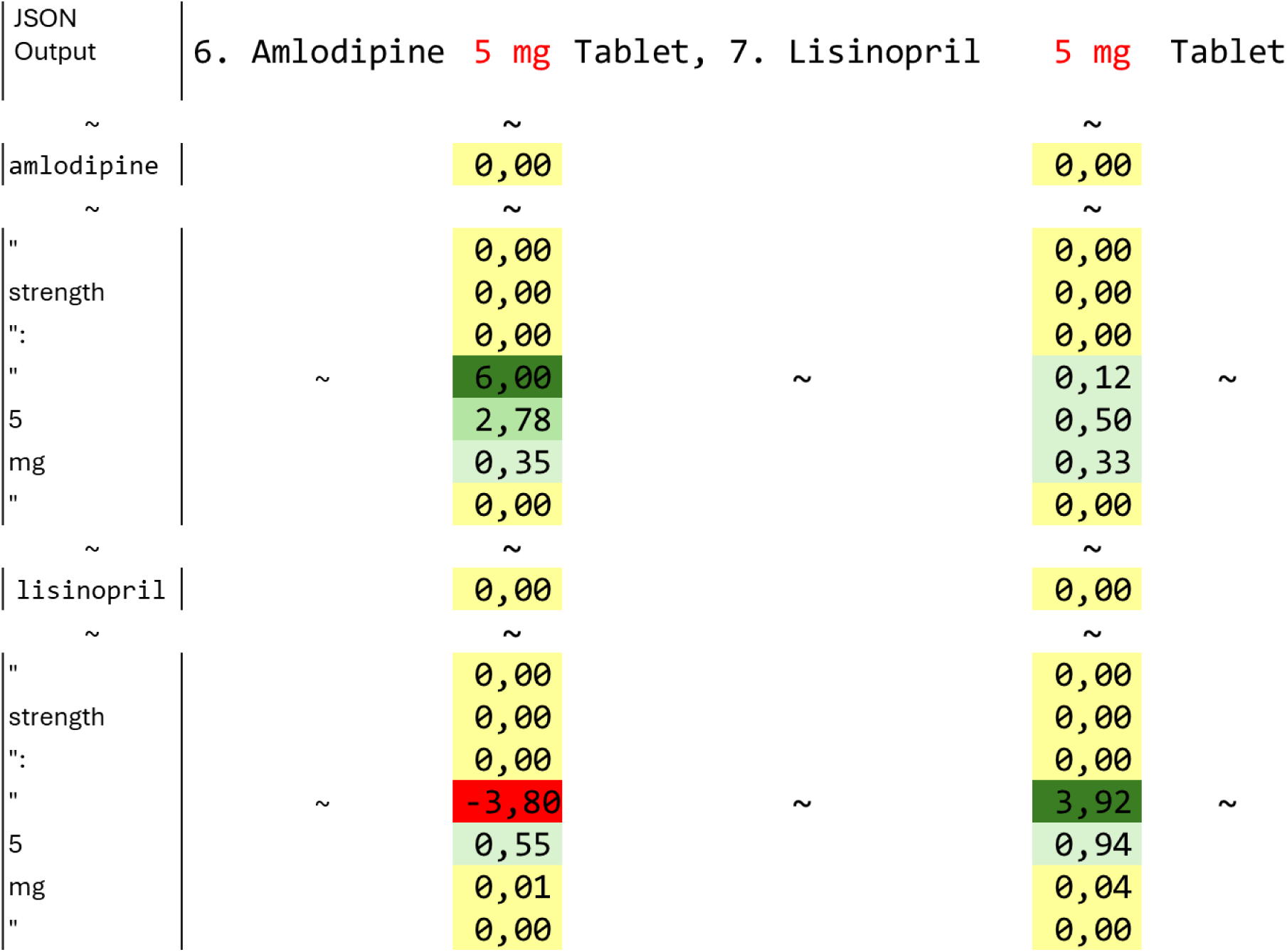
Visualizing approximate Shapley values for the strength token of the input text “6. Amlodipine 5 mg Tablet, 7. Lisinopril 5 mg Tablet” and the generated JSON token for the strength relation class and value.

**Suppl. Figure 3.**
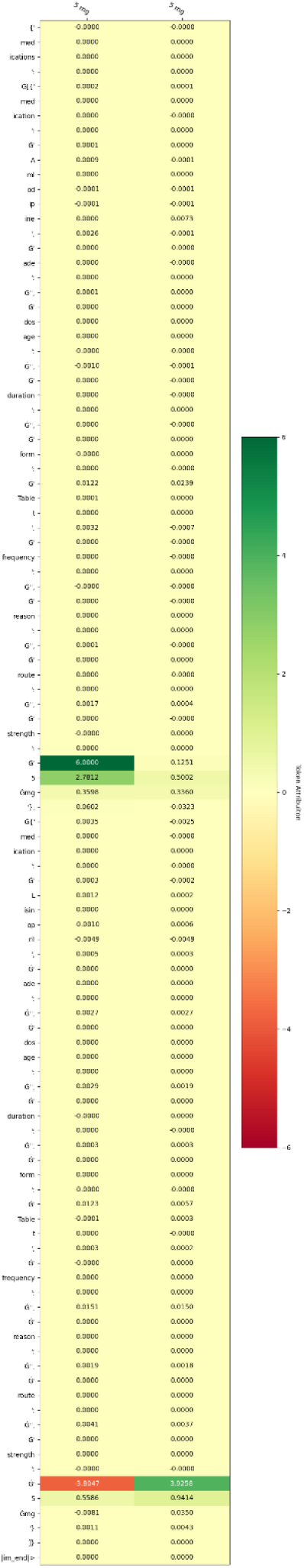
Example ellipsis 1: complete set of Shapley values for selected input token (strength values). Model: Llama 70b FT. Input text: “… 6. Amlodipine 5 mg Tablet, 7. Lisinopril 5 mg Tablet …”

##### Example 2

**Suppl. Figure 4.**
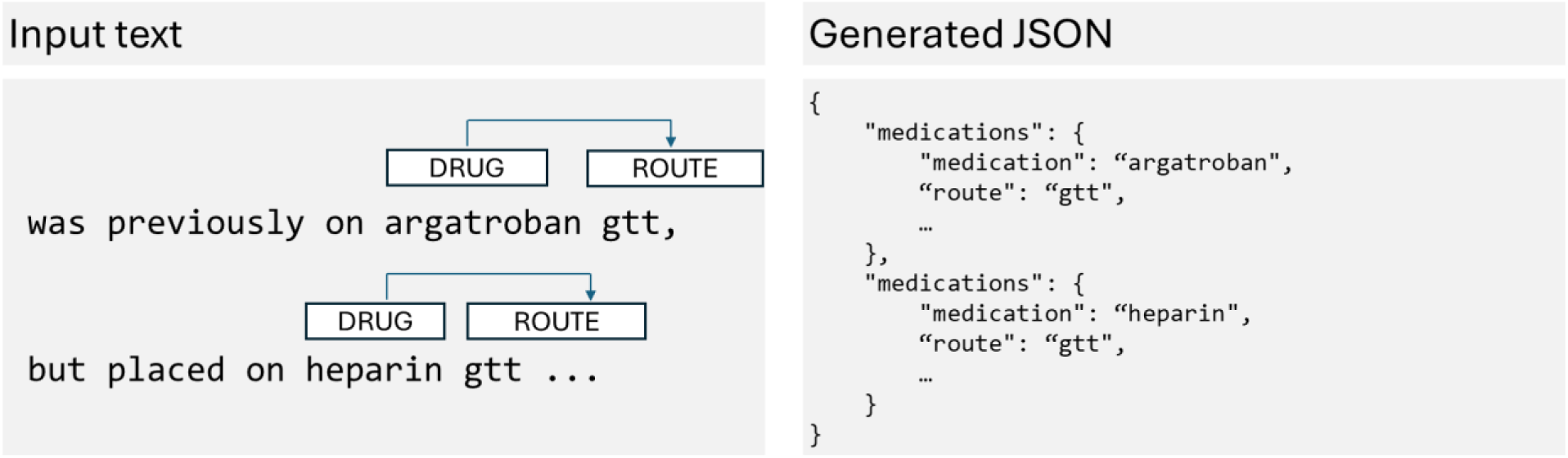
(left) Example input text containing two medications (Argatroban, Heparin) each related to a similar route value (gtt). (right) Corresponding generated JSON output snippet containing the medication names and the route values.

**Suppl. Figure 5.**
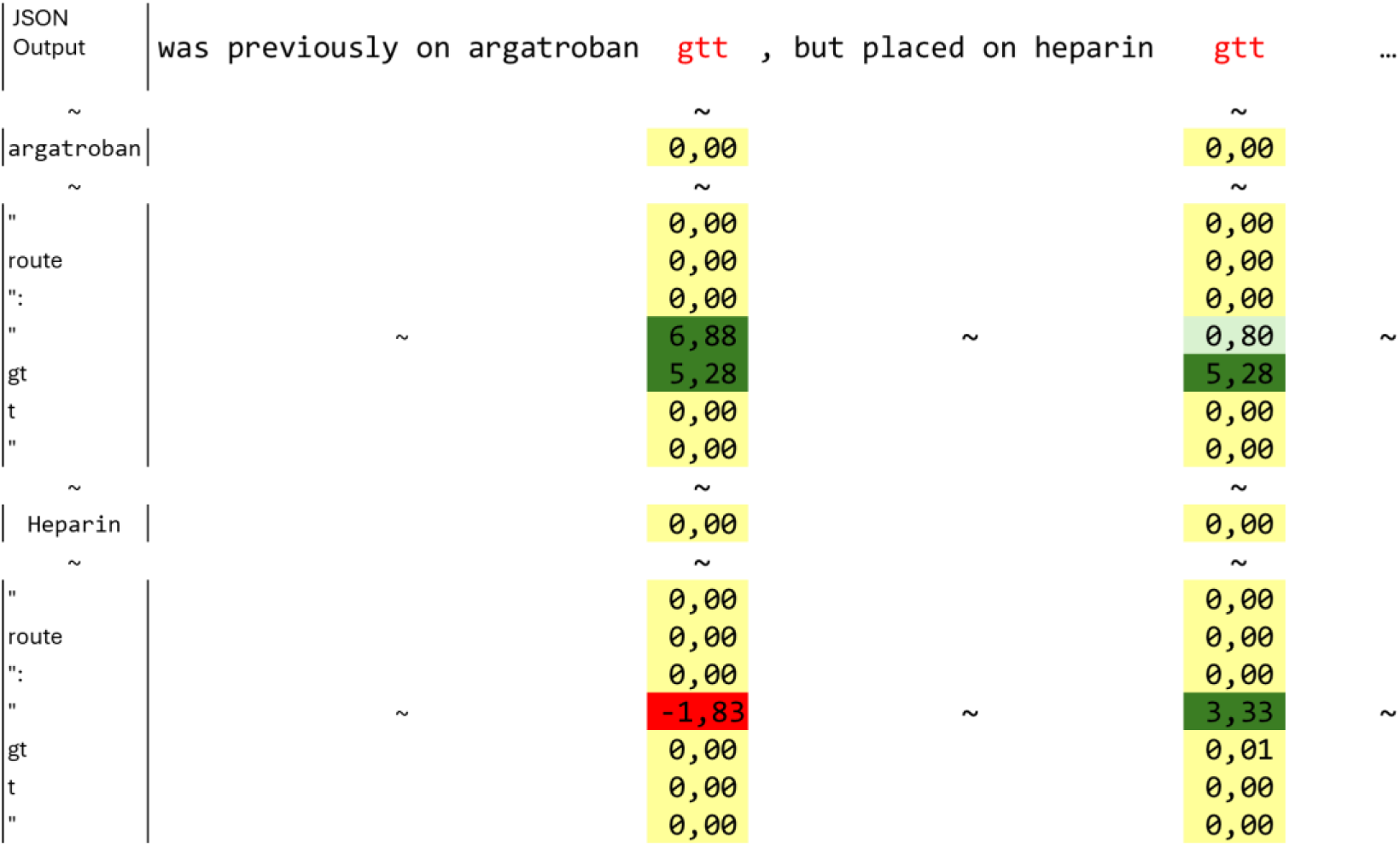
Visualizing approximate Shapley values for the route token of the input text “was previously on argatroban gtt, but placed on heparin gtt …” and the generated JSON token for the route relation class and value.

**Suppl. Figure 6.**
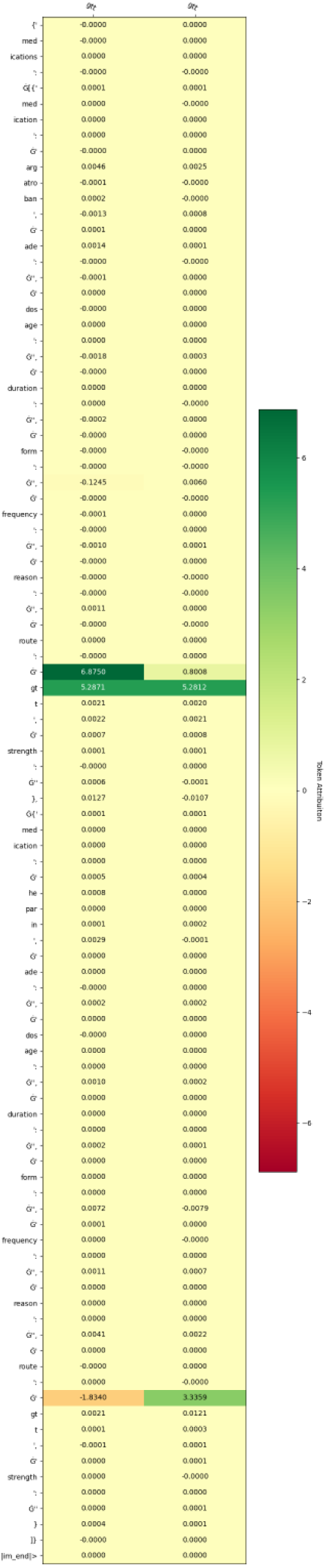
Example ellipsis 2: complete set of Shapley values for selected input token (route values). Model: Llama 70b FT. Input text: “… was previously on argatroban gtt, but placed on heparin gtt …”

##### Example 3

**Suppl. Figure 7.**
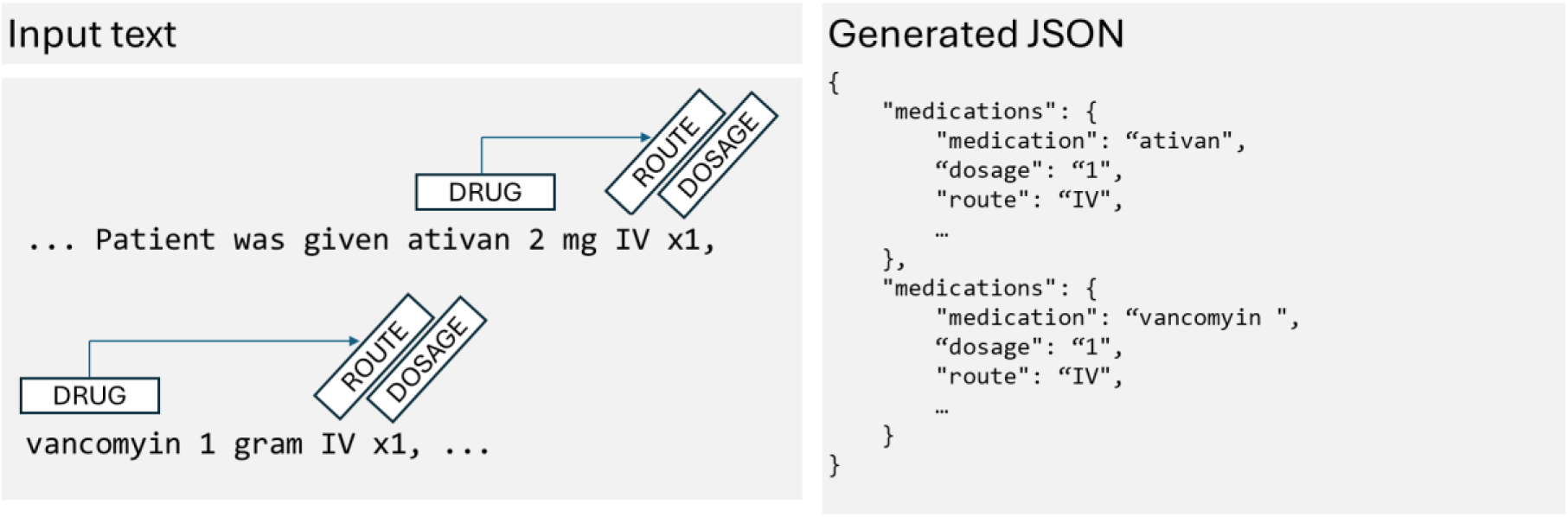
(left) Example input text containing two medications (Ativan, Vancomyin) each related to a similar route value (IV) and dosage value (1). (right) Corresponding generated JSON output snippet containing the medication names and the route and dosage values.

**Suppl. Figure 8.**
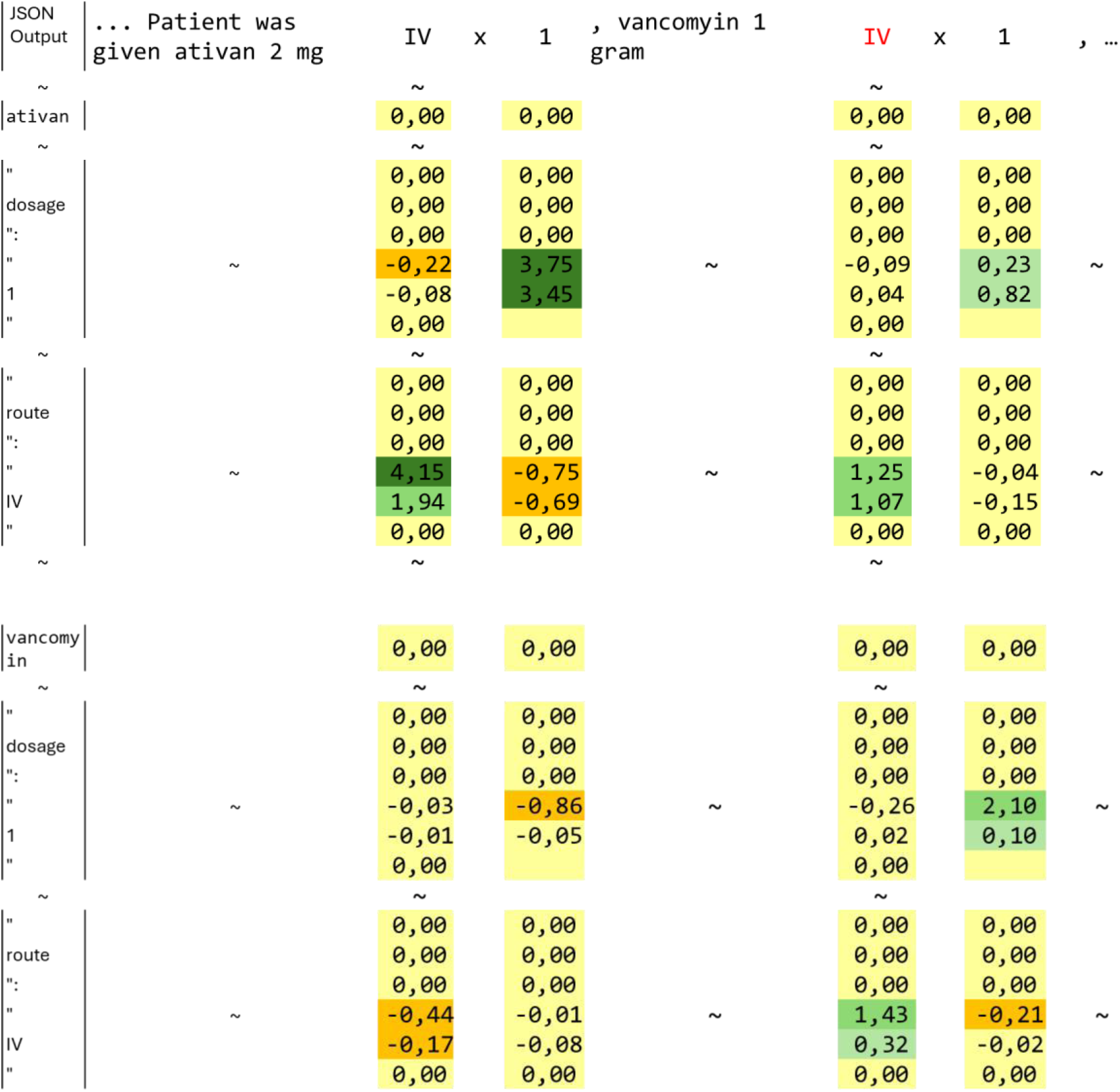
Visualizing approximate Shapley values for the dosage and route token of the input text “… Patient was given ativan 2 mg IV x1, vancomyin 1 gram IV x1, … “and the generated JSON token for the dosage and route relation class and value.

**Suppl. Figure 9.**
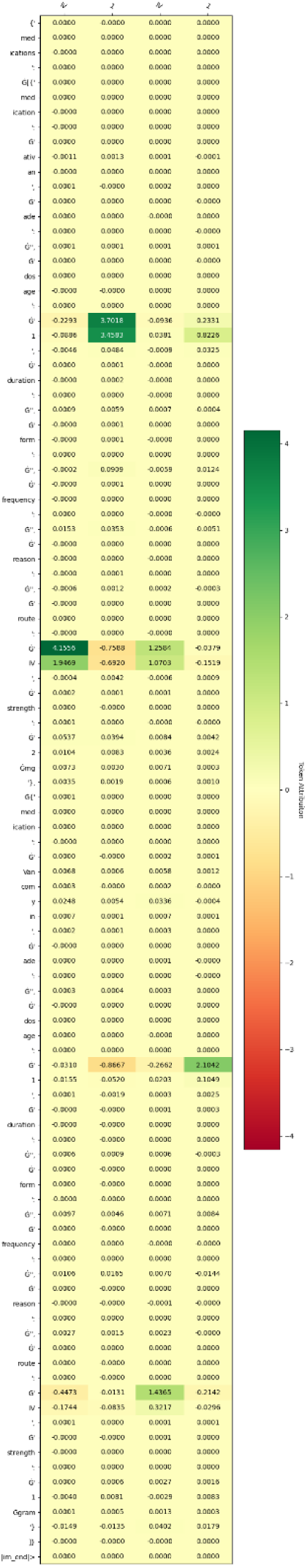
Example ellipsis 3: complete set of Shapley values for selected input token (dosage and route values). Model: Llama 8b FT. Input text: “… Patient was given ativan 2 mg IV x1, Vancomyin 1 gram IV x1, …”

#### Use case 2

The second use case uncovers that LLMs possess implicit knowledge regarding relation information, even when they do not explicitly generate this information in the JSON output. The following examples emphasize, that token containing information on adverse drug events or medication reason negatively contribute to an empty string generated as a relation value in the JSON output.

##### Example 1

**Suppl. Figure 10.**
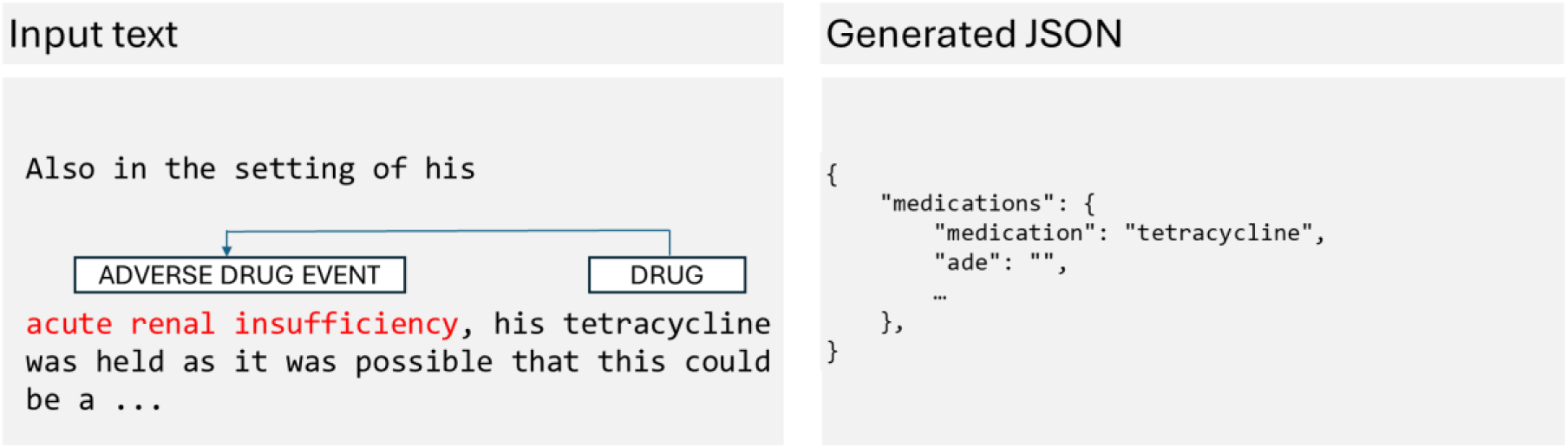
(left) Example input text containing ADE of the medication Tetracycline. (right) Corresponding generated JSON output snippet containing the medication name and the empty ADE value.

**Suppl. Figure 11.**
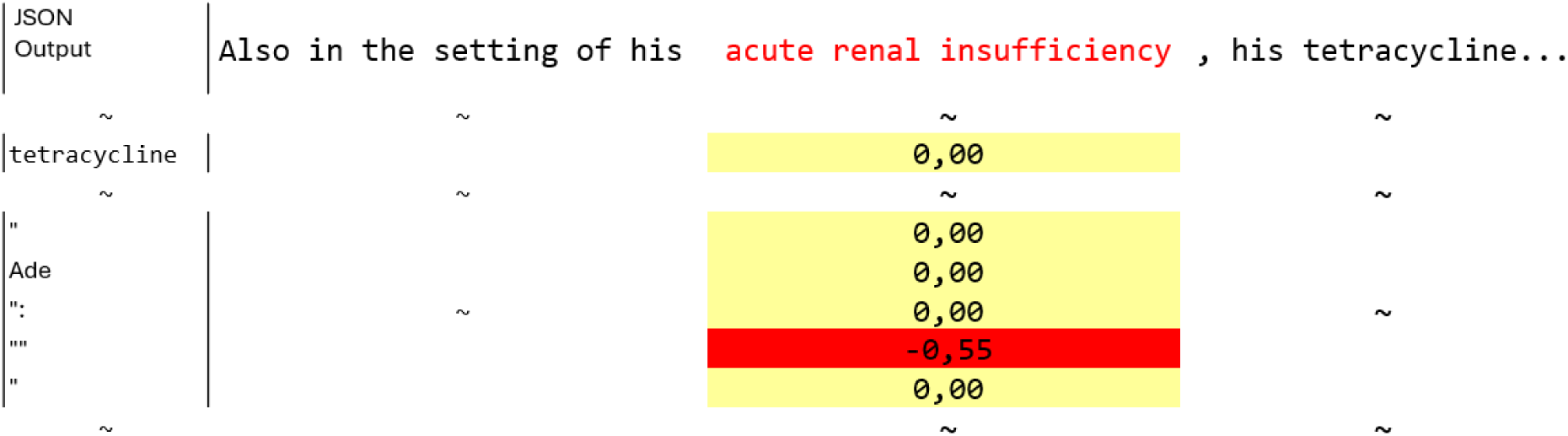
Visualizing approximate Shapley values for the ADE token of the input text “Also in the setting of his acute renal insufficiency, his tetracycline was held as it was possible that this could be a …” and the generated JSON token for the ADE relation class and value.

**Suppl. Figure 12.**
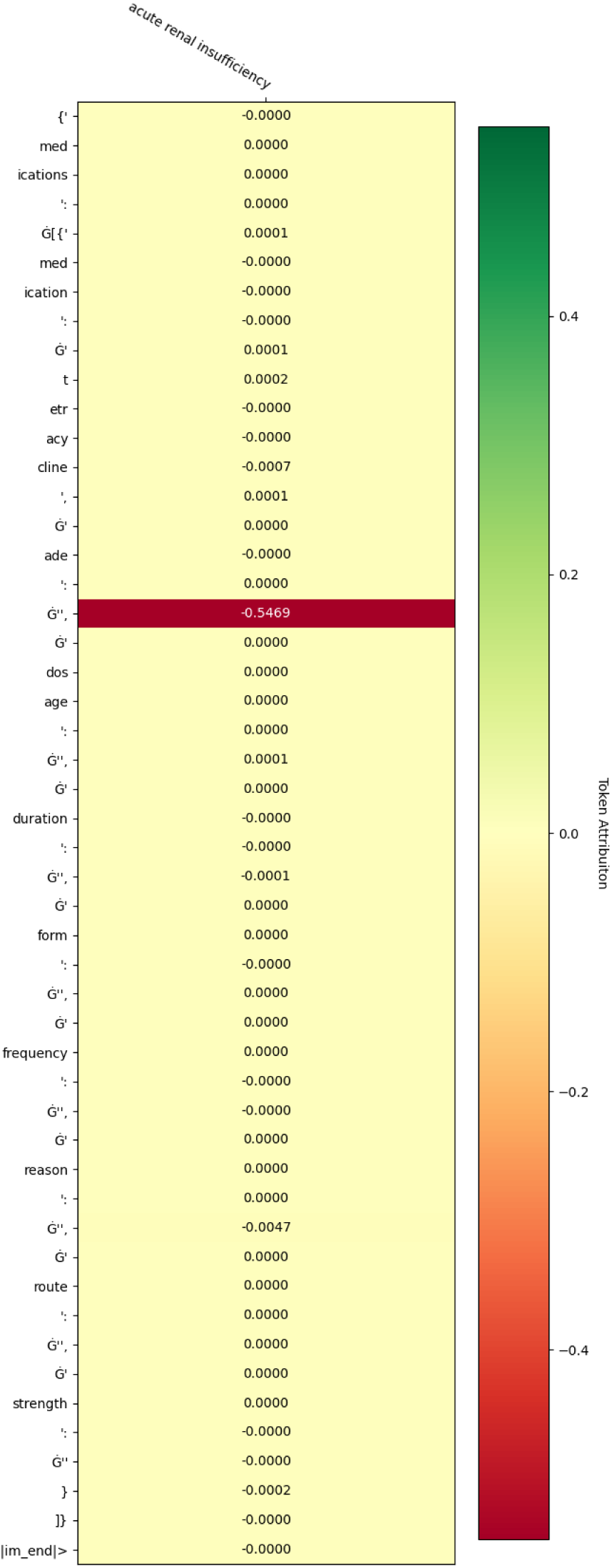
Example uncover 1: complete set of Shapley values for selected input token (ADE values). Model: Llama 8b FT. Input text: … Also in the setting of his acute renal insufficiency, his tetracycline was held as it was possible that this could be a …

##### Example 2

**Suppl. Figure 13.**
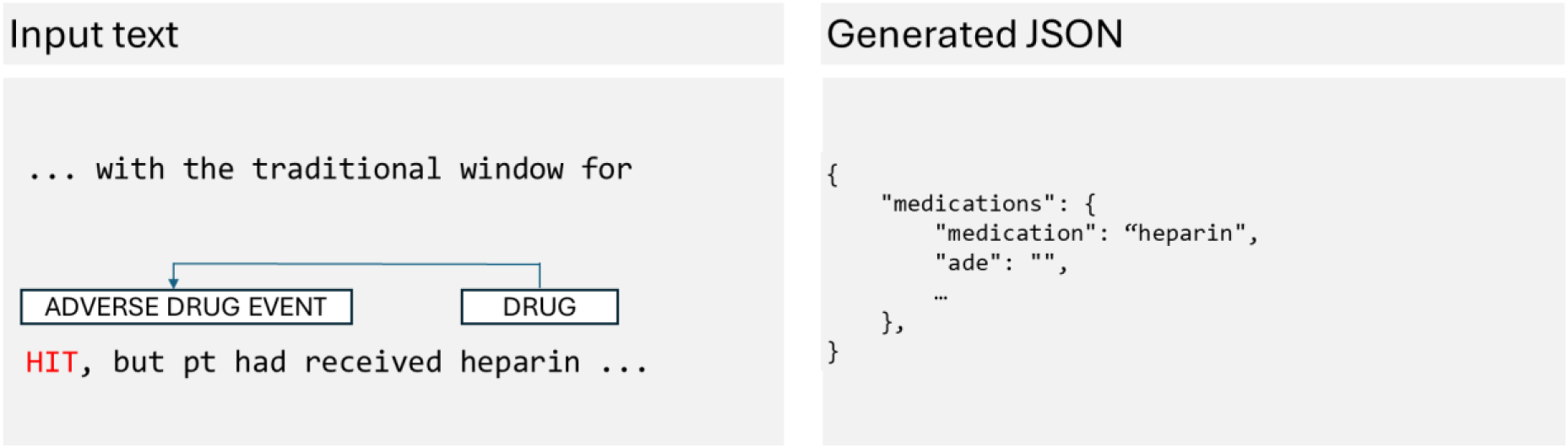
(left) Example input text containing ADE of the medication Heparin. (right) Corresponding generated JSON output snippet containing the medication name and the empty ADE value.

**Suppl. Figure 14.**
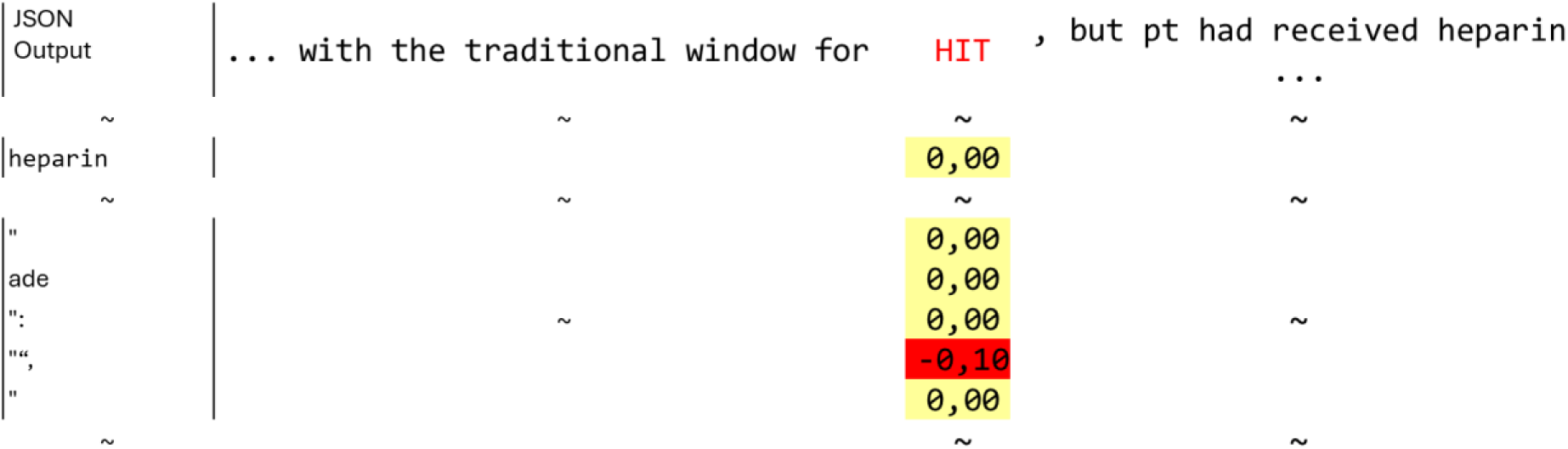
Visualizing approximate Shapley values for the ADE token of the input text “… with the traditional window for HIT, but pt had received heparin …” and the generated JSON token for the ADE relation class and value.

**Suppl. Figure 15.**
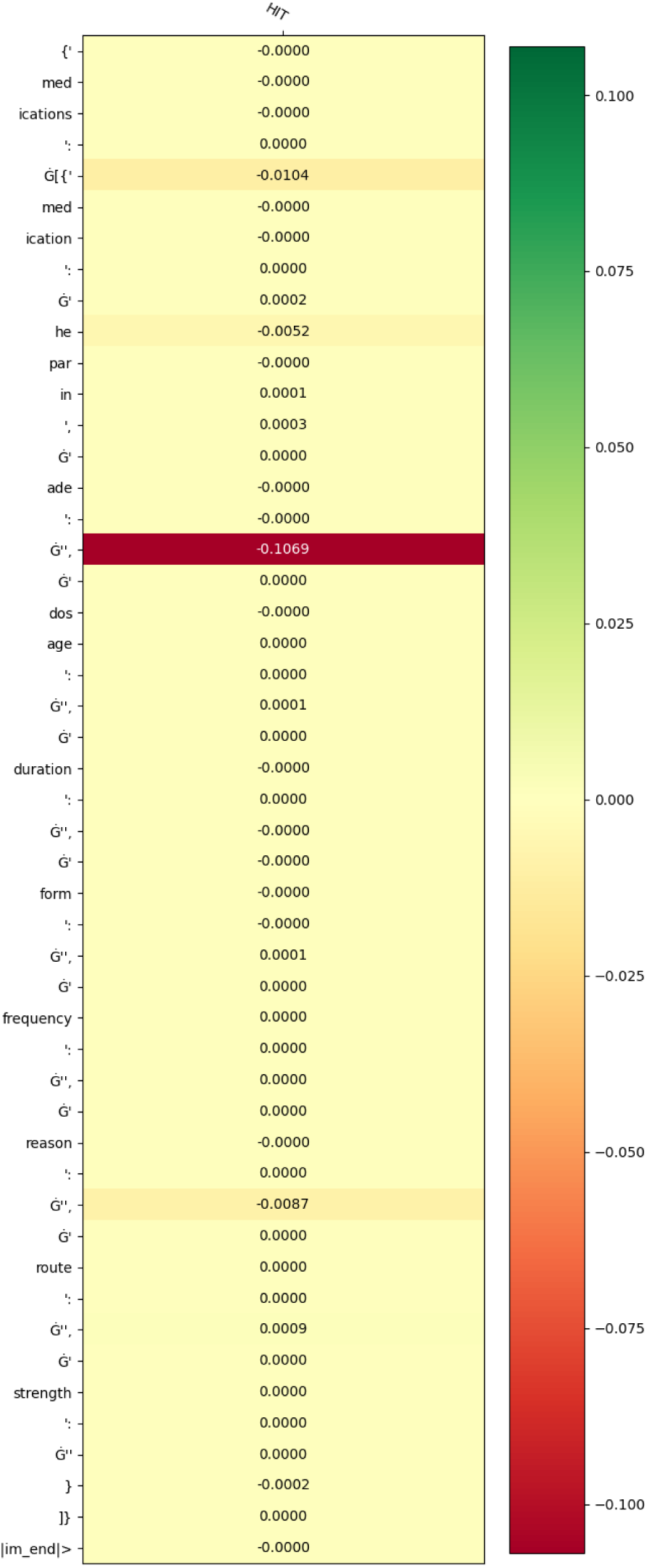
Example uncover 2: complete set of Shapley values for selected input token (ade value). Model: Llama 8b FT. Input text: … with the traditional window for HIT, but pt had received heparin …

##### Example 3

**Suppl. Figure 16.**
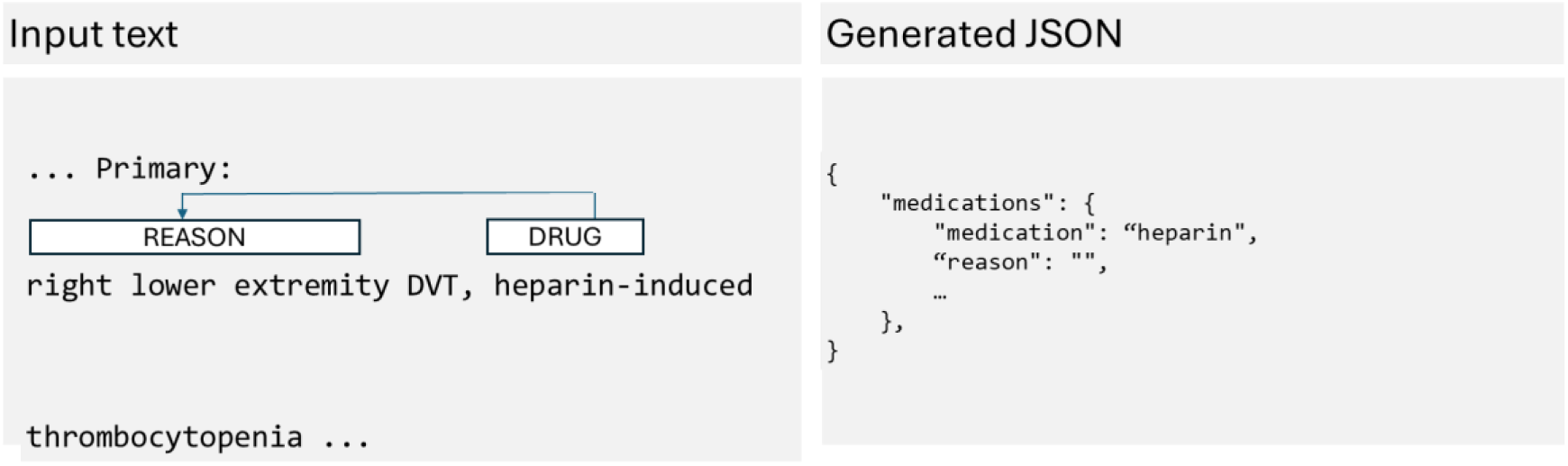
(left) Example input text containing reason of the medication heparin. (right) Corresponding generated JSON output snippet containing the medication name and the empty reason value.

**Suppl. Figure 17.**
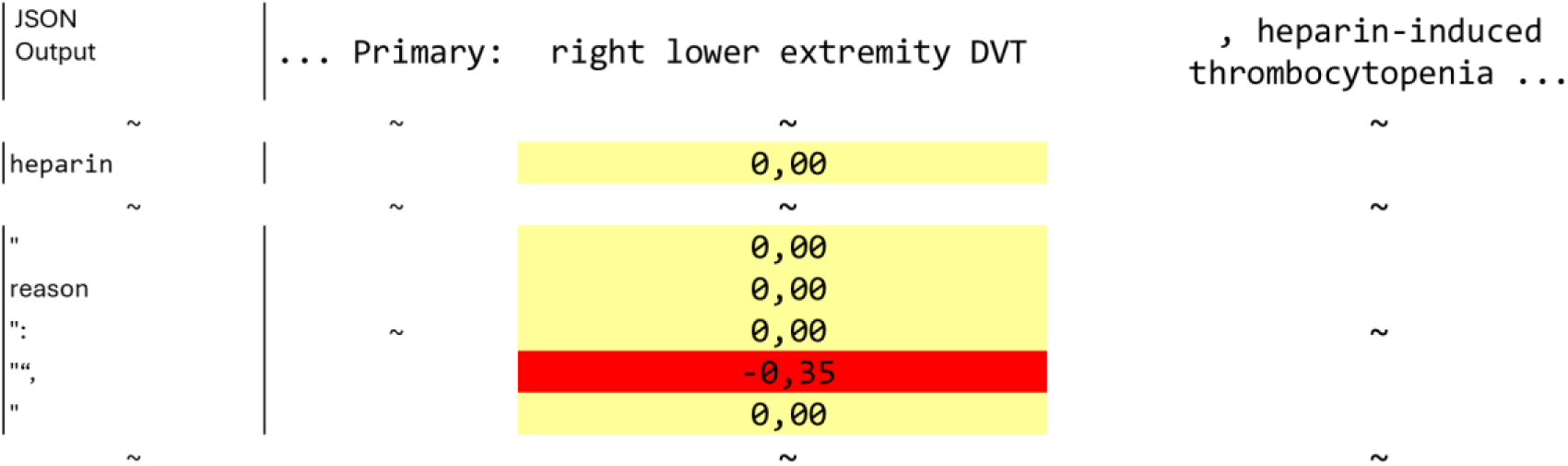
Visualizing approximate Shapley values for the reason token of the input text “… Primary: right lower extremity DVT, heparin-induced thrombocytopenia …” and the generated JSON token for the reason relation class and value.

**Suppl. Figure 18.**
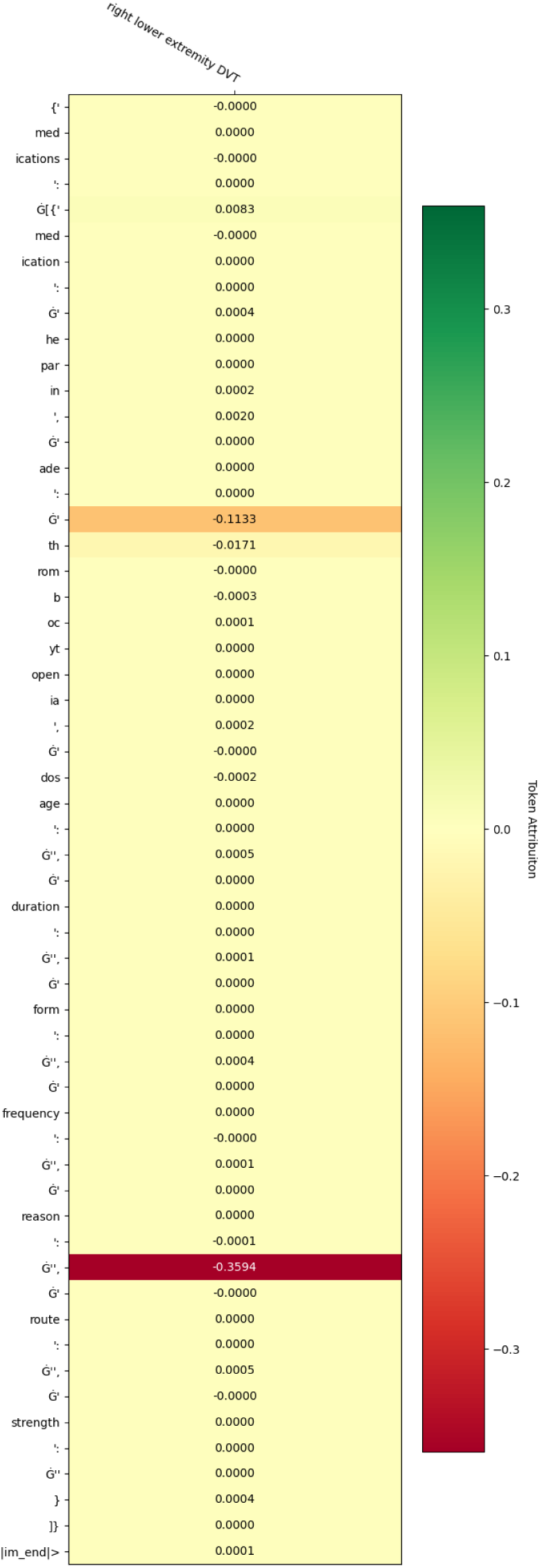
Example uncover 3: complete set of Shapley values for selected input token (reason value). Model: Llama 8b FT. Input text: … Primary: right lower extremity DVT, heparin-induced … … thrombocytopenia …

